# Patient-Level Risk Characterization of Drug-Associated Hidradenitis Suppurativa Using Machine Learning

**DOI:** 10.64898/2026.05.04.26352179

**Authors:** Kyle Maas, Claire Brewer, Aaron Chai, Dodie Park, Michelle Martin-Pozo, Elizabeth Phillips, Eric Mukherjee

## Abstract

**Importance:** Hidradenitis suppurativa (HS) has been reported with several medications, but the spectrum of implicated drug classes and patient-level risk factors remains poorly characterized.

**Objective:** To characterize drug-associated HS pharmacovigilance signals across drug classes, distinguish new-onset from worsened disease, and evaluate whether machine learning (ML) approaches identify reproducible patient-level predictors.

**Design:** Disproportionality and ML of spontaneous adverse event reports submitted from 2004 through 2023.

**Setting:** The US Food and Drug Administration Adverse Event Reporting System.

**Participants:** Reports containing a preferred term for hidradenitis, stratified by whether HS was a listed indication (worsened) or an adverse event only (new-onset). A parallel cohort comprised all tumor necrosis factor inhibitor (TNFi)-exposed reports.

**Exposures:** Primary suspect and concomitant medications.

**Main Outcomes and Measures:** Disproportionate reporting quantified by the reporting odds ratio; agent- and indication-specific hidradenitis suppurativa reporting rates; and discrimination, calibration, feature concordance, and risk enrichment of supervised machine-learning models estimating the probability of HS.

**Results:** Among 5529 HS reports (3725 with and 1804 without an indication), patients were predominantly female (3511 [63.5%]), with a mean (SD) age of 41 (14) years. Among reports without a hidradenitis suppurativa indication, significant signals included adalimumab (reporting odds ratio, 12.6; 95% CI, 11.3-14.0), infliximab (8.2; 6.7-9.9), secukinumab (6.6; 5.2-8.2), and isotretinoin (6.2; 4.2-8.9). Among TNFi exposed reports, reporting rates were highest for adalimumab (1.08 per 1000; OR, 2.75; 95% CI, 2.41-3.14) and lowest for etanercept (0.11 per 1000; OR, 0.12; 95% CI, 0.09-0.15), and the association was strongest in inflammatory bowel disease (OR 3.06; 2.20-4.25); all reported associations were significant at P < .001. Supervised ML models demonstrated similar discrimination while identifying consistent patient-level predictors of HS. Tree-based models showed greater enrichment of high-risk reports than penalized regression.

**Conclusions and Relevance:** HS reporting signals spanned biologic, hormonal, retinoid, and immunomodulatory drug classes. Multiple supervised ML approaches consistently identified TNFi agent (adalimumab), age, and a history of arthropathy as associated with paradoxical HS, suggesting that pharmacovigilance data contain reproducible information on heterogeneity of HS reporting. These findings support indication-aware monitoring when initiating these therapies, while recognizing that spontaneous reports reflect disproportionate reporting rather than incidence or causality.

**KEY POINTS:** *Question:* Which medications are associated with disproportionate reporting of hidradenitis suppurativa (HS), and can routinely collected pharmacovigilance data identify patient- and treatment-level characteristics associated with paradoxical HS?

*Findings:* In this disproportionality and machine-learning analysis of 5529 HS reports, significant reporting signals spanned tumor necrosis factor inhibitors, interleukin-17 inhibitors, retinoids, hormonal agents, and antineoplastics and were strongest for adalimumab and in inflammatory bowel disease. Calibrated risk models were concordant, and the best-performing model (gradient-boosted trees) enriched the highest-risk 1% approximately 16-fold for cases.

*Meaning:* Findings support indication-aware dermatologic monitoring when initiating implicated therapies, but reflect disproportionate reporting rather than incidence or causality.

## INTRODUCTION

Hidradenitis suppurativa (HS) is a chronic, inflammatory skin disorder that substantially affects quality of life through pain and disfigurement.^1^ Although HS usually arises spontaneously, case reports and series suggest that biologics, hormonal therapies, psychotropics, and gamma-secretase inhibitors may trigger new-onset disease or worsen pre-existing HS.^2–9^ Existing evidence, however, is limited to case reports and small series, leaving the broader landscape of drug-associated HS poorly characterized.

The paradoxical occurrence of HS during treatment with TNF-α inhibitors (TNFi) has received attention, with systematic reviews commonly implicating adalimumab and infliximab.^2,10,11^ Proposed mechanisms include compensatory upregulation of interferon-γ, unmasking of IL-17 driven neutrophilic inflammation, and plasmacytoid dendritic cell activation by type I interferons.^7,12–14^ Recently, paradoxical HS has been reported with IL-17 inhibitors (IL-17i), JAK inhibitors (JAKi), and integrin inhibitors, raising the possibility that drug-associated HS reporting varies across mechanisms, treatment indications, and clinical phenotypes.^2,7,15,16^

Rare paradoxical adverse events present two broad challenges. First, their low frequency limits characterization beyond conventional disproportionality analyses – even when association is recognized, small counts limit subgroup analysis. Second, even when an association is recognized, little is known about which patient and treatment characteristics are associated with development of these events.^17,18^ Most applications of machine learning (ML) in pharmacovigilance have focused on improving signal detection. In contrast, once a rare adverse event has been recognized, an equally important question is whether pharmacovigilance data can characterize the patient- and treatment-level features associated with that event.

The US Food and Drug Administration (FDA) Adverse Event Reporting System (FAERS), which contains >25 million reports, provides sufficient scale to study heterogeneity in reported rare events.^19,20^ We analyzed FAERS reports from 2004 through 2023 to quantify drug-specific reporting signals, distinguish new-onset from worsened disease, and evaluate variation across drugs and treatment indications. We compared multiple ML approaches to determine whether nonlinear models improved risk enrichment beyond regression, and whether patient-level risk could be estimated reproducibly across modeling strategies.^21–26^ Because FAERS is a spontaneous reporting system, these findings represent disproportionate reporting patterns rather than estimates of incidence, absolute risk, or causality.

## METHODS

### Study Design and Data Source

We conducted a retrospective pharmacovigilance study using deduplicated adverse event data from FAERS as previously described. ^27^ The overall study design is summarized in eFigure 1. Because the data were deidentified, institutional review board review was not required.

**Figure 1.**
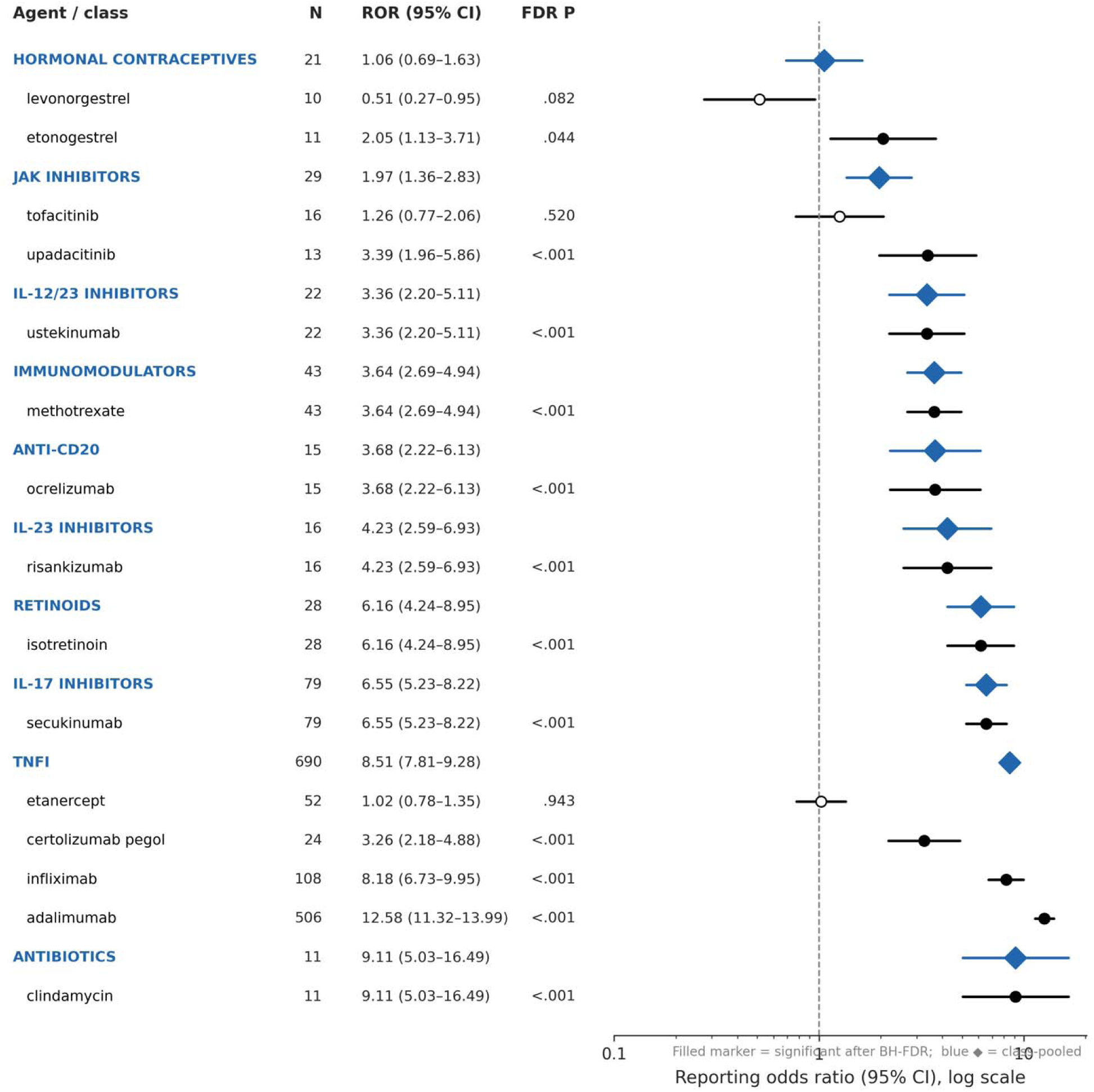
Forest Plot of Medication-Associated HS Rates by Agent (Drug-Induced HS) Forest plot of reporting odds ratios (ROR) with 95% CIs for drug–HS signals among reports without HS listed as a indication (drug-induced stratum). Drugs are grouped by pharmacologic class, with class-level pooled estimates shown in blue and individual agents shown in black. Only agents with at least 10 drug-induced HS reports are shown. N reports indicates HS case count for each agent or class. The x-axis is displayed on a log scale; the reference line at ROR = 1 corresponds to no disproportionality

### Case Identification, Stratification, and Disproportionality Analysis

Cohorts comprised reports containing a MedDRA preferred term including “hidradenitis” (eTable 1). Reports were stratified according to whether HS was listed as both an indication and an adverse event (HS-indication stratum) or solely as an adverse event (no-HS-indication stratum). For readability, these strata are subsequently referred to as potentially drug-worsened and potentially drug-associated new-onset HS, respectively. Standard disproportionality analyses including reporting odds ratios (ROR) were performed.

### Disease Context, Time-to-Onset, and Indication-Stratified Rate Analysis

Disease context was derived from the complete list of drug indications for each HS report, each defined by sets of Observational Medical Outcomes Partnership (OMOP) concept identifiers (eTable 1 in the Supplement). A single report may contribute to more than one disease-context category. Time-to-onset was calculated as the interval between event date (event_dt) and therapy start date, excluding missing, negative, or implausible (>50 years) values.

### Machine-Learning Cohort and Model Construction

To understand patient-level heterogeneity in drug-induced HS, we analyzed TNFi-exposed FAERS reports without an HS indication (n = 1 451 950). HS cases (n = 953; 0.07%) were defined by the MedDRA criteria above.

Predictors were selected a priori based on biological plausibility and/or availability within FAERS. Candidate predictors comprised demographics (age, sex, and missingness indicators), TNFi identity, indication-group flags (arthropathies, inflammatory bowel disease [IBD], psoriasis, and multiple indications), and concomitant medications present in >1% of reports (38 drugs), yielding 62 candidate features. Reports were split 70/30 with stratification on HS status (training, n = 1 016 365; test, n = 435 585), followed by feature selection used 10-fold cross-validated random forest (RF) permutation importance, retaining features selected in at least five folds. All feature selection and model fitting were confined to the training set; the held-out test set was used for final evaluation and model interpretation. Subsequently, we compared five supervised learning approaches representing complementary modeling paradigms: penalized linear models (elastic net [EN] and ridge logistic regression), tree-based ensembles (RF and XGBoost [XGB]), and an interpretable explainable boosting machine (EBM).^21–26^ Isolation Forest (IF) was included to test whether HS reports behaved as statistical outliers rather than a reproducible clinical phenotype.^28,29^

Because the objective was patient-level risk characterization rather than binary classification, model evaluation emphasized calibrated probability estimation and enrichment of higher-risk patients. Primary performance measures were the area under the precision-recall curve (AUPRC), interpreted relative to the no-skill baseline (equal to the event prevalence), and top-percentile risk enrichment. Calibration (Brier score and predicted-to-observed ratio) and discrimination (AUROC) were evaluated as secondary measures on the held-out test set.^30–32^ Model interpretation used algorithm-specific approaches, including permutation importance, TreeSHAP values, EBM shape functions and term importances, penalized-regression coefficients, and a SHAP-based interaction screen.^26,33–36^

An exploratory logistic regression evaluated interaction between treatment indication and TNFi agent with HS as the outcome, adjusting for age group, sex, and the 15 concomitant medications (selected to avoid overfitting and for computational tractability) with the highest feature importance in the supervised learning analyses. Cells with ≤4 HS cases were flagged as sparse because resulting estimates were considered unstable. The 20 interaction terms with sufficient cell counts (of 30 total) were corrected by Benjamini-Hochberg false discovery rate (FDR). Marginally standardized predicted probabilities were calculated for each indication × TNFi agent combination to facilitate interpretation of interaction effects.^37–39^

A full outline of methods, including class imbalance, model calibration, benchmarking, software, and specific parameters can be found in the eText.

## RESULTS

### Study Population

A total of 5529 reports of HS were identified, comprising 3725 drug-worsened and 1804 drug-induced reports (Table 1). Reports predominantly involved female patients (3511 [63.5%]), with a mean (SD) age of 41 (14) and similar demographics between strata; age was missing in 3205 (58.0%) reports. The US accounted for 3047 of 3725 drug-worsened reports (81.8%) but only 968 of 1804 drug-induced reports (53.7%). Relative to drug-worsened reports, drug-induced reports had higher proportions of hospitalization (409 [22.7%] vs 590 [15.8%]), other serious outcomes (1033 [57.3%] vs 1406 [37.7%]), and disability (54 [3.0%] vs 44 [1.2%]) (Table 1). Annual report counts and the age distribution are shown in eFigure 2.

**Figure 2.**
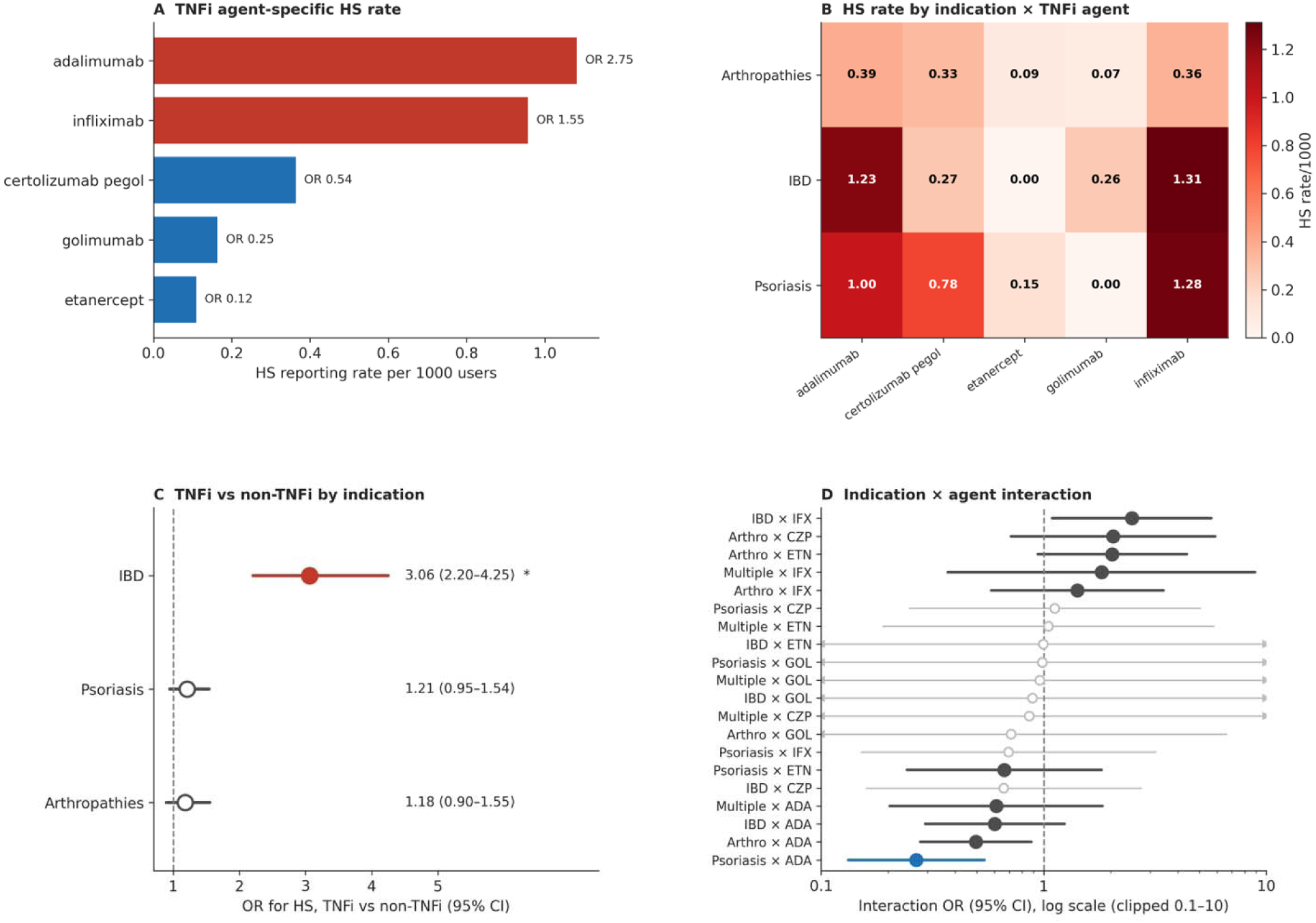
Tumor necrosis factor inhibitor (TNFi) agent-specific hidradenitis suppurativa (HS) reporting rates and indication × agent interactions. A, HS reporting rate per 1000 reports by TNFi agent, with odds ratio (OR) versus all other TNFi combine (adalimumab, 1.08, OR 2.75; infliximab, 0.96, OR 1.55; certolizumab pegol, 0.37, OR 0.54; golimumab, 0.17, OR 0.25; etanercept, 0.11, OR 0.12). B, HS reporting rate per 1000 by indication and TNFi agent. C, HS reporting rate per 1000 for TNFi versus non-TNFi users within each indication; the association was strongest in inflammator bowel disease (IBD) (OR, 3.06; P < .001), with nonsignificant differences in arthropathies (OR, 1.18) an psoriasis (OR, 1.21). D, Adjusted indication × TNFi-agent interaction odds ratios with 95% CIs from the logistic interaction model (adjusted for age group, sex, and 15 concomitant-drug covariates; 20 interaction terms, Benjamini-Hochberg corrected). Asterisk indicates raw P < .05; double asterisk, FDR P < .05 (psoriasis × adalimumab, OR 0.26); dagger, sparse cell (≤4 HS cases).

**Table 1.**
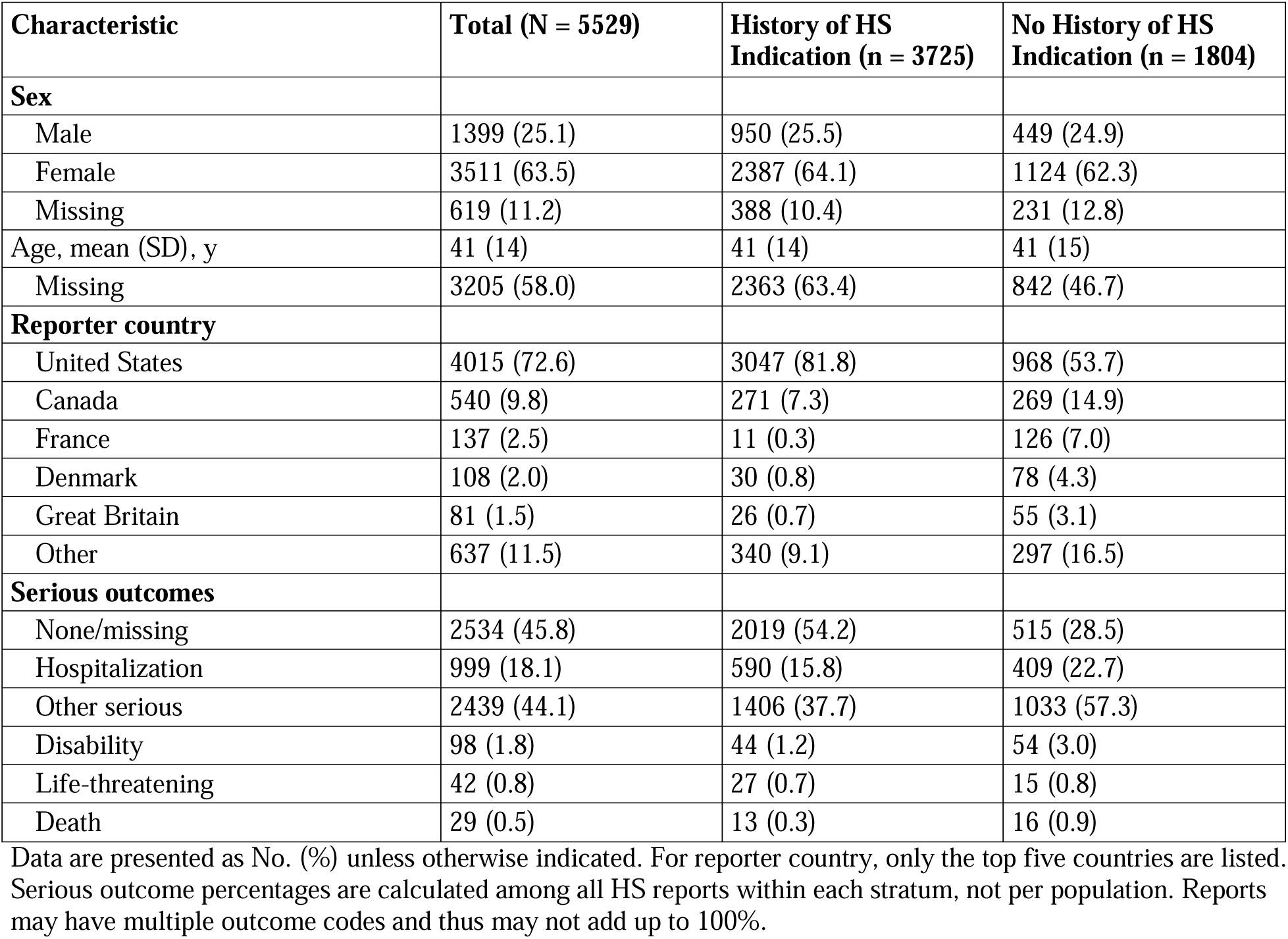
Demographic and Report Characteristics of HS Outcome Reports.

| **Characteristic** | **Total (N = 5529)** | **History of HS Indication (n = 3725)** | **No History of HS Indication (n = 1804)** |
| --- | --- | --- | --- |
| **Sex** |  |  |  |
| Male | 1399 (25.1) | 950 (25.5) | 449 (24.9) |
| Female | 3511 (63.5) | 2387 (64.1) | 1124 (62.3) |
| Missing | 619 (11.2) | 388 (10.4) | 231 (12.8) |
| Age, mean (SD), y | 41 (14) | 41 (14) | 41 (15) |
| Missing | 3205 (58.0) | 2363 (63.4) | 842 (46.7) |
| **Reporter country** |  |  |  |
| United States | 4015 (72.6) | 3047 (81.8) | 968 (53.7) |
| Canada | 540 (9.8) | 271 (7.3) | 269 (14.9) |
| France | 137 (2.5) | 11 (0.3) | 126 (7.0) |
| Denmark | 108 (2.0) | 30 (0.8) | 78 (4.3) |
| Great Britain | 81 (1.5) | 26 (0.7) | 55 (3.1) |
| Other | 637 (11.5) | 340 (9.1) | 297 (16.5) |
| **Serious outcomes** |  |  |  |
| None/missing | 2534 (45.8) | 2019 (54.2) | 515 (28.5) |
| Hospitalization | 999 (18.1) | 590 (15.8) | 409 (22.7) |
| Other serious | 2439 (44.1) | 1406 (37.7) | 1033 (57.3) |
| Disability | 98 (1.8) | 44 (1.2) | 54 (3.0) |
| Life-threatening | 42 (0.8) | 27 (0.7) | 15 (0.8) |
| Death | 29 (0.5) | 13 (0.3) | 16 (0.9) |

Data are presented as No. (%) unless otherwise indicated. For reporter country, only the top five countries are listed. Serious outcome percentages are calculated among all HS reports within each stratum, not per population. Reports may have multiple outcome codes and thus may not add up to 100%.

### Reported Indications and Disease Context

The reported indication for the primary suspect drug differed substantially by prior HS history. Among reports with a documented HS indication, HS itself overwhelmingly dominated, whereas among reports without an HS indication the leading indications were Crohn disease, psoriasis, and rheumatoid arthritis (eFigure 3). Indication-derived disease-context profiles likewise varied by primary suspect drug and by HS history (eFigure 4): IBD and arthropathy indications predominated for TNFi and integrin inhibitors, psoriasis clustered with IL-17i, and drug-worsened reports showed a higher prevalence of follicular-occlusion indications.

**Figure 3.**
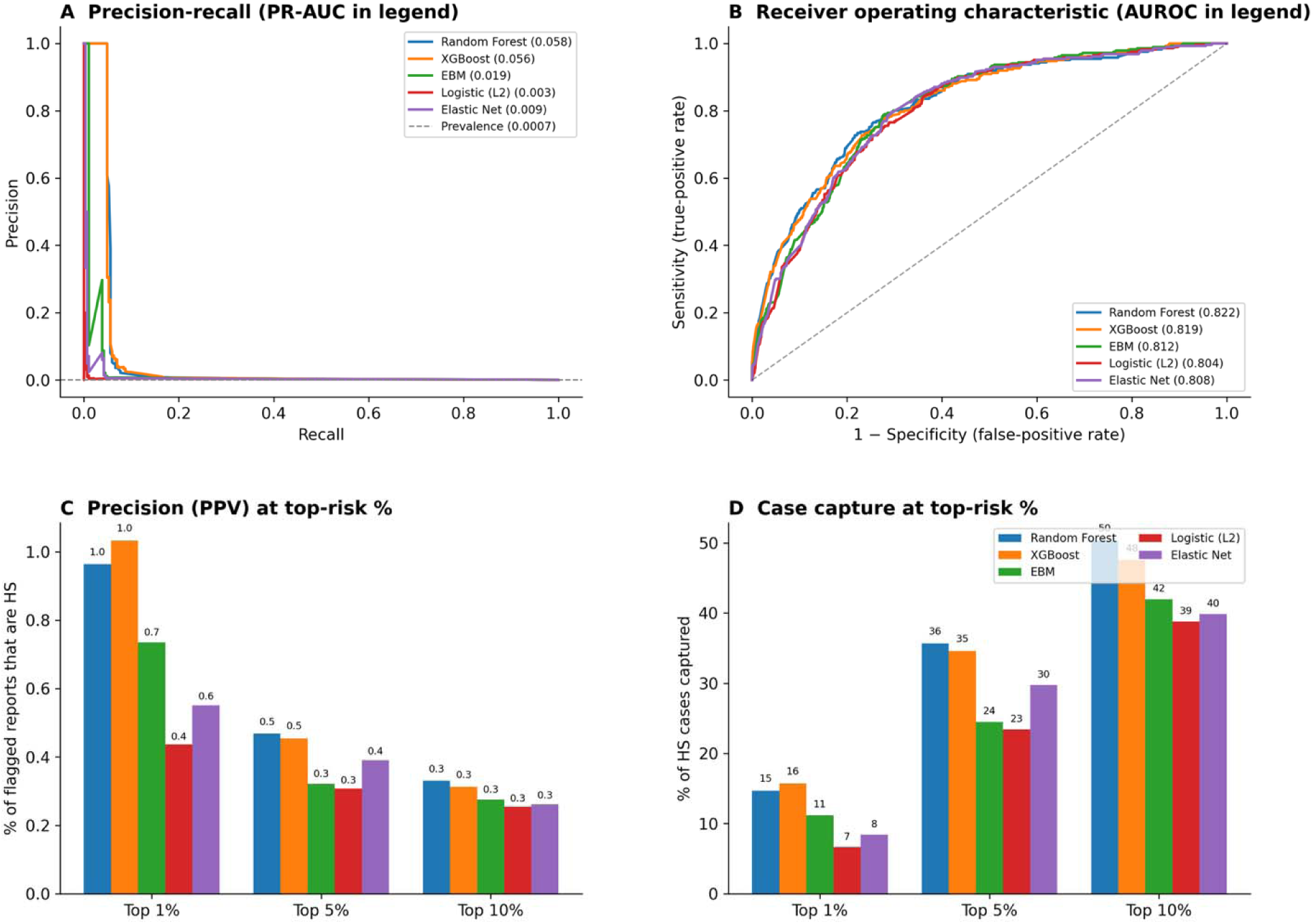
Machine-learning model performance and benchmark comparison (TNFi-exposed test set). Random forest, gradient-boosted trees (XGBoost), an explainable boosting machine, and penalized (ridge an elastic-net) logistic regression compared on the held-out test set (n = 435 585; 286 HS cases); an unsupervised isolation forest is shown as a label-free baseline. A, Precision-recall curves (area under the precision-recall curve in legend). B, Receiver operating characteristic curves (area under the curve in legend). C, Precision (positive predictive value) among the highest-risk 1%, 5%, and 10% of reports, by model. D, Case capture (percentage of all HS cases) within the same top-risk groups. Precision-recall and enrichment were emphasized because AUROC ma appear favorable despite limited positive predictive value at a 0.07% event proportion. Model calibration is shown in eFigure 9.

**Figure 4.**
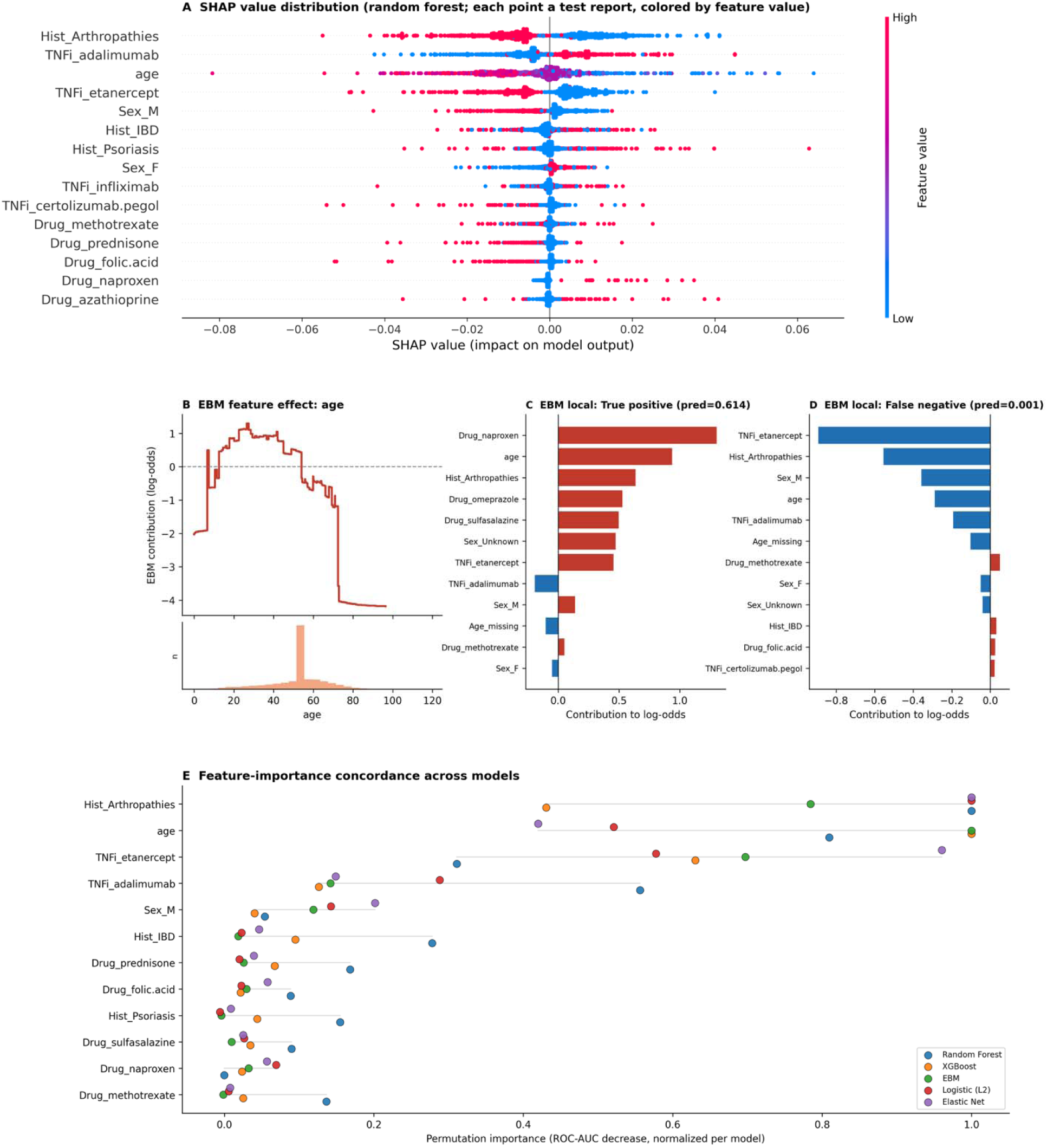
Predictors of tumor necrosis factor inhibitor (TNFi)-associated hidradenitis suppurativa (HS): Model interpretation and cross-model concordance. A, SHAP (Shapley additive explanations) value distribution for the leading features of the random forest; each point is a held-out test report, positioned by its SHAP value and colored by feature value. B, Explainable boostin machine (EBM) shape function for age (per-value contribution to the log-odds of HS reporting), with the ag distribution shown below. C and D, EBM local explanations for a correctly ranked true-positive report (C) and a missed false-negative report (D), showing each feature’s contribution to the predicted log-odds. E, Cross-model concordance of feature importance: permutation importance (decrease in the area under the receiver operatin characteristic curve when each feature is shuffled), normalized within each model, with one point per model. Reporting-completeness indicators (age and sex missingness) were included in all fitted models but are not shown.

### Disproportionality Signals

Significant disproportionality signals for HS were identified across multiple therapeutic classes and spanned both strata (eFigure 5; eTable 2). Adalimumab accounted for the largest HS report burden overall and showed prominent signals in both strata. TNFi as a class were the most consistently represented, with additional signals for infliximab, etanercept, certolizumab pegol, and golimumab. IL-17i, including secukinumab, also showed significant signals, as did emerging signals for JAK inhibitors, integrin inhibitors (vedolizumab, natalizumab), and the anti-CD20 agent ocrelizumab. Non-biologic signals included isotretinoin, hormonal agents (testosterone, etonogestrel), and antineoplastic agents (cytarabine, omacetaxine). Among drug-induced reports, class-level pooled signals were most prominent for TNFi and IL-17i, with substantial within-class variation across agents (Figure 1; eTable 3). After Benjamini-Hochberg correction within the drug-induced stratum, 34 of 83 signals remained significant (eTable 2).

Among drug-induced (no HS history) reports, time-to-onset varied across drug classes (eFigure 6; eTable 4). Among usable reports, TNFi showed gradual onset (median, 231 days; n = 98), whereas IL-17i showed rapid onset (median, 25 days; n = 17), with intermediate latency for JAK inhibitors (28 days), anti-CD20 agents (282 days), and checkpoint inhibitors (336 days). Sex-stratified signals showed some disparities: testosterone had one of the strongest signals among reports coded as female, whereas isotretinoin, leflunomide, and sorafenib were among the strongest male signals (eFigure 7; eTable 5). Of 111 sex-stratified signals, 48 remained significant after Benjamini-Hochberg correction within each sex-by-history stratum.

### HS Reporting Proportions Differ among TNFi

Within the TNFi-exposed population, agent-specific HS reporting proportions varied widely (Figure 2A; eTable 6). Adalimumab had the highest (1.08 per 1000 reports; OR, 2.75 vs other TNFi; 95% CI, 2.41-3.14; P < .001) and infliximab the second highest (0.96 per 1000; OR, 1.55; 95% CI, 1.31-1.84; P < .001), whereas golimumab (0.17 per 1000; OR, 0.25; 95% CI, 0.12-0.52; P < .001) and etanercept (0.11 per 1000; OR, 0.12; 95% CI, 0.09-0.15; P < .001) were lowest. Rates also differed across indications (Figure 2B). When TNFi-exposed reports were compared with non-TNFi reports within each indication (Figure 2C; eTable 7), the association was strongest in IBD, where TNFi exposure carried 3.06-fold higher HS reporting (95% CI, 2.20-4.25; P < .001); associations in arthropathies (OR, 1.18; 95% CI, 0.90-1.55; P = .25) and psoriasis (OR, 1.21; 95% CI, 0.95-1.54; P = .13) were not significant.

### Indication × TNFi-Agent Interactions

In a logistic model incorporating indication × TNFi-agent interaction terms and adjusting for age, sex, and 15 concomitant medications, adalimumab was associated with increased reporting odds (OR, 2.33) and etanercept was associated with lower odds (OR, 0.16); male sex was associated with lower odds (OR, 0.61) (eTable 8). Across the indication × agent grid (Figure 2D; eTable 9), most interaction OR were consistent with the independent main effects. Ten of 30 indication-by-agent cells contained 4 or fewer HS cases and yielded unstable estimates (eTable 10). Raw-significant interactions included arthropathies × adalimumab (OR, 0.51) and IBD × infliximab (OR, 2.49), but after Benjamini-Hochberg correction across 20 interaction terms only psoriasis × adalimumab remained significant (OR, 0.26; 95% CI, 0.13-0.53; FDR P = .004), indicating lower HS reporting than expected from the separate associations of psoriasis and adalimumab (eTable 11).

### ML Models Enrich HS Reporting Among TNFi-Exposed Reports

We compared five supervised learning models and an unsupervised Isolation Forest to characterize patient-level risk of HS among TNFi-exposed reports using 29 of 62 candidate features (eTable 12). In the held-out test set (n = 435 585; 286 HS cases), supervised models showed similar discrimination (AUROC, 0.82-0.83), but precision-recall performance (AUPRC) and top-percentile enrichment differed by model. RF and XGB had the highest AUPRCs (0.05 each), compared with the prevalence of 0.0007, followed by EBM (0.03) and ridge regression (0.01) (Figure 3A and 3B; eTable 13). IF performed near chance (AUROC, 0.55; AUPRC, 0.001; eFigure 8).

After isotonic scaling, predicted-to-observed ratios were approximately 1.0 across supervised models (eFigure 9; eTables 13-17). Because the models estimated patient-level risk rather than producing binary classifications, we next examined their ability to enrich higher-risk patients. XGB enriched the top 1%, 5%, and 10% of reports 15.7-, 7.3-, and 5.3-fold, capturing 15.7%, 36.4%, and 53.1% of cases. RF captured 14.3%, 34.3%, and 50.3%, while penalized models had lower AUPRC and case capture at the same top-risk thresholds (Figure 3C and 3D; eTable 13). Given their comparable discrimination and shared leading predictors, we illustrate model interpretation using the calibrated random forest (the prespecified reference model; eTable 18) and the explainable boosting machine (Figure 4).

### ML Models of TNFi-Associated HS Show Concordance of Feature Importance

The random forest, EBM, and penalized regression models identified a broadly overlapping set of leading features, including arthropathy indication, age, TNFi identity, and IBD indication (Figure 4A). The EBM age function showed a nonlinear association, with higher model contributions across much of early and middle adulthood and lower contributions at older ages (Figure 4B). Local EBM explanations for a correctly ranked (true-positive) versus a missed (false-negative) report illustrated how the same features combine differently at the individual level (Figure 4C, 4D). The direction and magnitude of several concomitant-medication associations were corroborated by the logistic regression, the primary inferential model (eTable 19), and permutation importance (eTable 20). A complementary SHAP-based screen of pairwise feature interactions is provided in eTable 21. Across the five supervised models, feature-importance rankings showed substantial but nonuniform agreement (Figure 4E; eFigure 10; eTable 22). Pairwise top-10 overlap averaged 7.1 features, and 6 features appeared among the top 10 in all 5 models. Concordance was highest among EBM, ridge regression, and elastic net, while RF and XGB also showed similar rankings.

## DISCUSSION

In this large pharmacovigilance study, drug-associated HS reporting varied across medication classes, treatment indications, sex strata, and reported time to onset. These patterns likely reflect a combination of treatment context, underlying disease, patient susceptibility, diagnostic overlap, and reporting behavior. Among all medications, TNFi were the most consistently represented class, with adalimumab and infliximab contributing the strongest signals and etanercept showing the lowest reporting proportion. Consistent with an indication-dependent profile, drug-induced HS reports most commonly involved primary suspect drugs prescribed for Crohn’s disease, psoriasis, or rheumatoid arthritis, with the strongest indication-specific association observed in IBD.^40–43^ This association may partly reflect diagnostic overlap with perianal Crohn disease, although shared inflammatory susceptibility and treatment selection may also contribute.^44^ A leading proposed mechanism for TNFi-associated HS is an altered balance among inflammatory cytokines that may unmask IL-17, IL-1β, and neutrophil-driven follicular inflammation in predisposed patients.^12,16^

Because HS was reported in only 0.07% of TNFi-exposed reports. At this frequency, FAERS-based models are not suitable for actionable individual prediction, but can be used to characterize relative reporting probability and enrichment across exposed reports. Rather than identifying individual patients destined to develop HS, our models estimated relative reporting risk across the TNFi-exposed population. Although overall discrimination was similar across supervised ML approaches, all concentrated a disproportionate share of HS reports into high-risk strata: the highest-ranked 1% of reports was enriched ∼16-fold relative to the background event rate of the full TNFi-exposed population. The near-chance performance of IF suggests that HS reports were not identifiable as nonspecific outliers; discrimination required supervised learning from labeled combinations of patient-, indication-, and treatment-level features.^17,29,45^

Interestingly, despite their fundamentally different assumptions, five supervised learning models agreed on an average of 7.1 of 10 top predictors, suggesting the associations reflect stable clinical structure rather than technical artifacts.^23,45^ This pattern suggests a stable core of patient- and treatment-level features (particularly age, TNFi identity, and a history of arthropathy), with differences in how modeling approaches prioritized the remaining predictors. Agreement was strongest among EBM and the penalized logistic-regression models, while RF and XGB also showed similar rankings; XGB had intermediate concordance with both groups.

Several concomitant medication associations among TNFi recipients (eg, naproxen) may also reflect confounding by indication, as patients with more severe, painful disease are both more likely to receive these medications and more likely to develop HS, or protopathic bias, where treatment was initiated for symptoms of undiagnosed HS.^46^ The single interaction surviving correction, a decreased-risk psoriasis × adalimumab interaction, should be interpreted cautiously; it may reflect distinct cytokine axes in psoriasis that mitigate adalimumab risk, though further study is required.^47^

Our analysis extended beyond TNFi. Reports involving IL-17i, particularly secukinumab, had a shorter median time-to-onset than TNFi, although this may reflect shorter follow-up and small case numbers. The paradox that IL-17 blockade may both treat established HS and be associated with new or worsening disease could reflect redundancy within the IL-17A/F axis or confounding by baseline indication.^7,48,49^ If confirmed in clinically adjudicated cohorts, the shorter reported latency could suggest differences in the timing or mechanism of HS emergence across biologic classes. Additional signals were generated from JAK inhibitors, integrin inhibitors, and anti-CD20 therapy, though interpretation should be cautious due to limited case counts, heterogeneous indications, and potential confounding by indication.^40,50–52^ The isotretinoin signal may reflect shared follicular-occlusion susceptibility, diagnostic overlap with severe acne, or treatment of prodromal HS symptoms rather than a direct drug effect.^53–55^ Testosterone and etonogestrel signals are consistent with the hormonal sensitivity of HS and show a strong sex-specific disproportionality.^56,57^ Signals from antineoplastics like cytarabine and omacetaxine should be interpreted with particular caution, as may partly represent neutrophilic eccrine hidradenitis or other HS-like eruptions rather than classic HS.^58–60^

## LIMITATIONS

Several limitations inherent to pharmacovigilance should be considered. As a spontaneous reporting system, FAERS cannot estimate incidence or establish causality, is subject to missing data and reporting bias.^61–65^ Confounding by indication is particularly important, because many implicated medications are prescribed for diseases independently associated with HS.^46^ Similarly, classification of reports as drug-induced vs drug-worsened depends on whether HS was listed as an indication and may result in misclassification. Time-to-onset analyses were limited by missing or inaccurate therapy dates, and disease context was inferred from reported indications rather than complete charts.

The ML models inherit these limitations – predicted probabilities should be interpreted as reporting risk within the FAERS population, rather that absolute risk. Model performance and calibration were internally evaluated in a held-out FAERS set and should not be assumed to generalize to clinical populations. In addition, important clinical variables, like smoking, disease severity, body mass index, etc., were unavailable and may lead to residual confounding. For interaction models, sparse indication-by-agent cells limited estimation. External validation in population-based cohorts or electronic health record datasets will be important to determine the generalizability of these patient-level risk patterns.

## CONCLUSION

This FAERS analysis identified heterogeneous HS reporting signals across multiple drug classes, most consistently TNF and IL-17 inhibitors. The findings suggest that routinely collected pharmacovigilance data contain reproducible patient- and treatment-level information that can be leveraged to characterize heterogeneity in rare paradoxical adverse events. By integrating disproportionality analysis, logistic regression with interaction terms, and ML, we extends pharmacovigilance beyond signal detection toward characterization of reporting-risk heterogeneity.^66,67^ Clinically, these findings support careful review of medication timing and indication when HS develops or unexpectedly worsens. Pharmacovigilance signals should inform clinical suspicion, indication-aware monitoring, and shared decision-making rather than discontinuation of an effective therapy. Prospective cohorts and clinically-adjudicated datasets are needed to validate these signals, distinguish true drug-induced HS from confounding by indication and HS-like eruptions, and determine whether the approach improves evaluation of other rare drug-event pairs.

## Ethics approval and consent to participate

This study used publicly available, de-identified adverse event reporting data from the FDA Adverse Event Reporting System (FAERS). Institutional review board approval and informed consent were not required.

## Consent for publication

Not applicable.

## Author approval

All authors have reviewed and approved the final version of the manuscript and agree to its submission.

## Data availability

Source FAERS data are publicly available from the FDA: https://www.fda.gov/drugs/fda-adverse-event-monitoring-system-aems/faers-quarterly-data-files-documentation. Analytic code and derived data sufficient to reproduce the analyses will be deposited at GitHub upon publication.

## Acknowledgements

None

## Patient Consent

Not applicable

## IRB Approval Status

Not applicable

## Use of AI

Generative artificial intelligence (Gemini 3.1 Pro, GPT-5.6) was used to assist with code development for statistical analyses, figure generation (though no figures were generated directly by AI), and editing; all code, outputs, and interpretation were reviewed and verified by the authors, who take full responsibility for the integrity and accuracy of the work.

## References

1. Sabat R, Alavi A, Wolk K, et al. Hidradenitis suppurativa. Lancet. 2025;405(10476):420–438. doi:10.1016/S0140-6736(24)02475-9

2. Frew JW, Vekic DA, Woods JA, Cains GD. Drug-associated hidradenitis suppurativa: A systematic review of case reports. Journal of the American Academy of Dermatology. 2018;78(1):217–219.e2. doi:10.1016/j.jaad.2017.08.046

3. Kisule A, Kak V, Alamelumangapuram C, Robinson C. Drug-Induced Hidradenitis Suppurativa: A Case Report. Cureus. 15(11):e49637. doi:10.7759/cureus.49637

4. Wut T, Vynnytska A, Ali A, Tiesenga F. Uncommon Drug-Induced Hidradenitis Suppurativa: A Case Report of A Patient on Lithium Therapy. Cureus. 16(10):e72049. doi:10.7759/cureus.72049

5. O’Sullivan Coyne G, Woodring TS, Lee CCR, Chen AP, Kong HH. Hidradenitis Suppurativa-Like Lesions Associated with Pharmacologic Inhibition of Gamma-Secretase. J Invest Dermatol. 2018;138(4):979–981. doi:10.1016/j.jid.2017.09.051

6. Machan A, Azendour H, Toufik H, Achemlal L, Boui M, Hjira N. Leflunomide-Induced Hidradenitis Suppurativa. Case Rep Rheumatol. 2020;2020:3549491. doi:10.1155/2020/3549491

7. Navarro-Triviño FJ, Sanchez-Parera R, Ruiz-Villaverde R. Secukinumab-induced paradoxical hidradenitis suppurativa. Dermatologic Therapy. 2020;33(1):e13150. doi:10.1111/dth.13150

8. Frantz TC, Kirwin D, Brahe C. Hidradenitis secondary to nirogacestat, a recently approved desmoid tumor medication. JAAD Case Rep. 2025;57:1–2. doi:10.1016/j.jdcr.2024.12.022

9. Riew GJ, Guggina LM, Repetto F, et al. Dermatologic adverse events associated with gamma secretase inhibitor nirogacestat: A retrospective multicenter cohort study. Journal of the American Academy of Dermatology. Published online August 2025:S0190962225025848. doi:10.1016/j.jaad.2025.08.002

10. Ward D, Nyboe Andersen N, Gørtz S, et al. Tumor Necrosis Factor Inhibitors in Inflammatory Bowel Disease and Risk of Immune Mediated Inflammatory Diseases. Clin Gastroenterol Hepatol. 2024;22(1):135–143.e8. doi:10.1016/j.cgh.2023.06.025

11. Murphy MJ, Cohen JM, Vesely MD, Damsky W. Paradoxical eruptions to targeted therapies in dermatology: A systematic review and analysis. Journal of the American Academy of Dermatology. 2022;86(5):1080–1091. doi:10.1016/j.jaad.2020.12.010

12. Behrangi E, Moodi F, Jafarzadeh A, Goodarzi A. Paradoxical and bimodal immune mediated dermatological side effects of TNF α inhibitors: A comprehensive review. Skin Res Technol. 2024;30(5):e13718. doi:10.1111/srt.13718

13. Salvador-Rodriguez L, Montero-Vílchez T, Arias-Santiago S, Molina-Leyva A. Paradoxical Hidradenitis Suppurativa in Patients Receiving TNF-α Inhibitors: Case Series, Systematic Review, and Case Meta-Analysis. Dermatology. 2020;236(4):307–313. doi:10.1159/000506074

14. De Stefano L, Pallavicini FB, Mauric E, et al. Tumor necrosis factor-α inhibitor-related autoimmune disorders. Autoimmun Rev. 2023;22(7):103332. doi:10.1016/j.autrev.2023.103332

15. Faivre C, Villani AP, Aubin F, et al. Hidradenitis suppurativa (HS): An unrecognized paradoxical effect of biologic agents (BA) used in chronic inflammatory diseases. J Am Acad Dermatol. 2016;74(6):1153–1159. doi:10.1016/j.jaad.2016.01.018

16. Salvatori S, Marafini I, Monteleone G. Paradoxical hidradenitis suppurativa in Crohn’s disease patients receiving infliximab: a case report and review of literature. Eur J Gastroenterol Hepatol. 2021;33(1S Suppl 1):e1046–e1050. doi:10.1097/MEG.0000000000002170

17. Arku D, Yousef C, Abraham I. Changing paradigms in detecting rare adverse drug reactions: from disproportionality analysis, old and new, to machine learning. Expert Opin Drug Saf. 2022;21(10):1235–1238. doi:10.1080/14740338.2022.2131770

18. Harpaz R, DuMouchel W, Shah NH, Madigan D, Ryan P, Friedman C. Novel Data-Mining Methodologies for Adverse Drug Event Discovery and Analysis. Clinical Pharmacology & Therapeutics. 2012;91(6):1010–1021. doi:10.1038/clpt.2012.50

19. Potter E, Reyes M, Naples J, Pan GD. FDA Adverse Event Reporting System (FAERS) Essentials: A Guide to Understanding, Applying, and Interpreting Adverse Event Data Reported to FAERS. Clin Pharmacol Ther. 2025;118(3):567–582. doi:10.1002/cpt.3701

20. Bate A, Evans SJW. Quantitative signal detection using spontaneous ADR reporting. Pharmacoepidemiology and Drug. 2009;18(6):427–436. doi:10.1002/pds.1742

21. Bae JH, Baek YH, Lee JE, Song I, Lee JH, Shin JY. Machine Learning for Detection of Safety Signals From Spontaneous Reporting System Data: Example of Nivolumab and Docetaxel. Front Pharmacol. 2020;11:602365. doi:10.3389/fphar.2020.602365

22. Lee JE, Kim JH, Bae JH, Song I, Shin JY. Detecting early safety signals of infliximab using machine learning algorithms in the Korea adverse event reporting system. Sci Rep. 2022;12(1):14869. doi:10.1038/s41598-022-18522-z

23. Chalabianloo N, Ahmadi F, Omrani MA, et al. Machine learning methods for predicting adverse drug events: A systematic review. Br J Clin Pharmacol. 2026;92(2):422–444. doi:10.1002/bcp.70377

24. Breiman L. Random Forests. Machine Learning. 2001;45(1):5–32. doi:10.1023/A:1010933404324

25. Chen T, Guestrin C. XGBoost: A Scalable Tree Boosting System. In: Proceedings of the 22nd ACM SIGKDD International Conference on Knowledge Discovery and Data Mining. KDD ’16. Association for Computing Machinery; 2016:785–794. doi:10.1145/2939672.2939785

26. Hegselmann S, Ertmer C, Volkert T, Gottschalk A, Dugas M, Varghese J. Development and validation of an interpretable 3 day intensive care unit readmission prediction model using explainable boosting machines. Front Med (Lausanne). 2022;9:960296. doi:10.3389/fmed.2022.960296

27. Mukherjee EM, Park D, Asiaee A, et al. Demographics, Overlap, and Latency of Severe Cutaneous Adverse Reactions in an FDA Database. medRxiv. Preprint posted online December 27, 2025:2025.03.05.25323441. doi:10.1101/2025.03.05.25323441

28. Liu FT, Ting KM, Zhou ZH. Isolation Forest. In: 2008 Eighth IEEE International Conference on Data Mining. 2008:413–422. doi:10.1109/ICDM.2008.17

29. Ricci C, Alberici L, D’Ambra V, et al. When survival models fail: An interpretable anomaly-detection approach for high-risk phenotypes in resected solid pseudopapillary tumors. Surgery. 2026;197:110381. doi:10.1016/j.surg.2026.110381

30. Saito T, Rehmsmeier M. The precision-recall plot is more informative than the ROC plot when evaluating binary classifiers on imbalanced datasets. PLoS One. 2015;10(3):e0118432. doi:10.1371/journal.pone.0118432

31. Ozenne B, Subtil F, Maucort-Boulch D. The precision--recall curve overcame the optimism of the receiver operating characteristic curve in rare diseases. J Clin Epidemiol. 2015;68(8):855–859. doi:10.1016/j.jclinepi.2015.02.010

32. Davis J, Goadrich M. The relationship between Precision-Recall and ROC curves. In: Proceedings of the 23rd International Conference on Machine Learning. ICML ’06. Association for Computing Machinery; 2006:233–240. doi:10.1145/1143844.1143874

33. Lundberg SM, Lee SI. A unified approach to interpreting model predictions. In: Proceedings of the 31st International Conference on Neural Information Processing Systems. NIPS’17. Curran Associates Inc.; 2017:4768–4777. Accessed July 20, 2026. https://dl.acm.org/doi/10.5555/3295222.3295230

34. Lou Y, Caruana R, Gehrke J, Hooker G. Accurate intelligible models with pairwise interactions. In: Proceedings of the 19th ACM SIGKDD International Conference on Knowledge Discovery and Data Mining. KDD ’13. Association for Computing Machinery; 2013:623–631. doi:10.1145/2487575.2487579

35. Ward IR, Wang L, Lu J, Bennamoun M, Dwivedi G, Sanfilippo FM. Explainable artificial intelligence for pharmacovigilance: What features are important when predicting adverse outcomes? Comput Methods Programs Biomed. 2021;212:106415. doi:10.1016/j.cmpb.2021.106415

36. Barus R, Schiro P, Faillie JL, et al. Applying machine learning to pharmacovigilance data: A proof-of-concept study. Br J Clin Pharmacol. Published online June 17, 2026. doi:10.1002/bcp.70655

37. Skrondal A. Interaction as departure from additivity in case-control studies: a cautionary note. Am J Epidemiol. 2003;158(3):251–258. doi:10.1093/aje/kwg113

38. Muller CJ, MacLehose RF. Estimating predicted probabilities from logistic regression: different methods correspond to different target populations. Int J Epidemiol. 2014;43(3):962–970. doi:10.1093/ije/dyu029

39. Benjamini Y, Hochberg Y. Controlling the False Discovery Rate: A Practical and Powerful Approach to Multiple Testing. Journal of the Royal Statistical Society Series B: Statistical Methodology. 1995;57(1):289–300. doi:10.1111/j.2517-6161.1995.tb02031.x

40. Brydges HT, Onuh OC, Friedman R, et al. Autoimmune, Autoinflammatory Disease and Cutaneous Malignancy Associations with Hidradenitis Suppurativa: A Cross-Sectional Study. Am J Clin Dermatol. 2024;25(3):473–484. doi:10.1007/s40257-024-00844-5

41. Garg A, Malviya N, Strunk A, et al. Comorbidity screening in hidradenitis suppurativa: Evidence-based recommendations from the US and Canadian Hidradenitis Suppurativa Foundations. J Am Acad Dermatol. 2022;86(5):1092–1101. doi:10.1016/j.jaad.2021.01.059

42. Maronese CA, Moltrasio C, Marzano AV. Hidradenitis Suppurativa-Related Autoinflammatory Syndromes: An Updated Review on the Clinics, Genetics, and Treatment of Pyoderma gangrenosum, Acne and Suppurative Hidradenitis (PASH), Pyogenic Arthritis, Pyoderma gangrenosum, Acne and Suppurative Hidradenitis (PAPASH), Synovitis, Acne, Pustulosis, Hyperostosis and Osteitis (SAPHO), and Rarer Forms. Dermatologic Clinics. 2024;42(2):247–265. doi:10.1016/j.det.2023.12.004

43. Garcovich S, Genovese G, Moltrasio C, Malvaso D, Marzano AV. PASH, PAPASH, PsAPASH, and PASS: The autoinflammatory syndromes of hidradenitis suppurativa. Clinics in Dermatology. 2021;39(2):240–247. doi:10.1016/j.clindermatol.2020.10.016

44. Amhis M, Belarbi KN, Bourrat E, Nassif A, Viala J, Martinez Vinson C. Differential Diagnosis Between Perianal Crohn’s Disease and Hidradenitis Suppurativa: A Challenging Teamwork. JPGN rep. 2021;2(2):e081. doi:10.1097/PG9.0000000000000081

45. Kompa B, Hakim JB, Palepu A, et al. Artificial Intelligence Based on Machine Learning in Pharmacovigilance: A Scoping Review. Drug Saf. 2022;45(5):477–491. doi:10.1007/s40264-022-01176-1

46. Sendor R, Stürmer T. Core concepts in pharmacoepidemiology: Confounding by indication and the role of active comparators. Pharmacoepidemiology and Drug. 2022;31(3):261–269. doi:10.1002/pds.5407

47. Armstrong AW, Read C. Pathophysiology, Clinical Presentation, and Treatment of Psoriasis: A Review. JAMA. 2020;323(19):1945–1960. doi:10.1001/jama.2020.4006

48. Fletcher JM, Moran B, Petrasca A, Smith CM. IL-17 in inflammatory skin diseases psoriasis and hidradenitis suppurativa. Clinical and Experimental Immunology. 2020;201(2):121–134. doi:10.1111/cei.13449

49. Melnik BC, John SM, Chen W, Plewig G. T helper 17 cell/regulatory T-cell imbalance in hidradenitis suppurativa/acne inversa: the link to hair follicle dissection, obesity, smoking and autoimmune comorbidities. Br J Dermatol. Published online June 11, 2018. doi:10.1111/bjd.16561

50. Hanzel J, Ma C, Casteele NV, Khanna R, Jairath V, Feagan BG. Vedolizumab and Extraintestinal Manifestations in Inflammatory Bowel Disease. Drugs. 2021;81(3):333–347. doi:10.1007/s40265-020-01460-3

51. Börnsen L, Christensen JR, Ratzer R, et al. Effect of Natalizumab on Circulating CD4+ T-Cells in Multiple Sclerosis. Oreja-Guevara C, ed. PLoS ONE. 2012;7(11):e47578. doi:10.1371/journal.pone.0047578

52. Kimura K, Nakamura M, Sato W, et al. Disrupted balance of T cells under natalizumab treatment in multiple sclerosis. Neurol Neuroimmunol Neuroinflamm. 2016;3(2):e210. doi:10.1212/NXI.0000000000000210

53. Daoud M, Suppa M, Heudens S, et al. Treatment of Acne with Isotretinoin Should Be Avoided in Patients with Hidradenitis Suppurativa “Conglobata Phenotype.” Dermatology. 2023;239(5):738–745. doi:10.1159/000530664

54. Poli F, Wolkenstein P, Revuz J. Back and Face Involvement in Hidradenitis Suppurativa. Dermatology. 2010;221(2):137–141. doi:10.1159/000315508

55. Desta B, Korytnikova E, Tessier-Kay M, et al. The role of γ-secretase in familial hidradenitis suppurativa: Implications for pathogenesis and targeted therapy. JAAD Case Rep. 2025;68:62–63. doi:10.1016/j.jdcr.2025.11.010

56. Grymowicz M, Rudnicka E, Podfigurna A, et al. Hormonal Effects on Hair Follicles. Int J Mol Sci. 2020;21(15):5342. doi:10.3390/ijms21155342

57. Abu Rached N, Gambichler T, Dietrich JW, et al. The Role of Hormones in Hidradenitis Suppurativa: A Systematic Review. Int J Mol Sci. 2022;23(23):15250. doi:10.3390/ijms232315250

58. Brehler R, Reimann S, Bonsmann G, Metze D. Neutrophilic Hidradenitis Induced by Chemotherapy Involves Eccrine and Apocrine Glands: The American Journal of Dermatopathology. 1997;19(1):73–78. doi:10.1097/00000372-199702000-00013

59. Flynn TC, Harrist TJ, Murphy GF, Loss RW, Moschella SL. Neutrophilic eccrine hidradenitis: A distinctive rash associated with cytarabine therapy and acute leukemia. Journal of the American Academy of Dermatology. 1984;11(4):584–590. doi:10.1016/S0190-9622(84)70210-6

60. Khoury HJ, Cortes J, Baccarani M, et al. Omacetaxine mepesuccinate in patients with advanced chronic myeloid leukemia with resistance or intolerance to tyrosine kinase inhibitors. Leukemia & Lymphoma. 2015;56(1):120–127. doi:10.3109/10428194.2014.889826

61. Alatawi YM, Hansen RA. Empirical estimation of under-reporting in the U.S. Food and Drug Administration Adverse Event Reporting System (FAERS). Expert Opin Drug Saf. 2017;16(7):761–767. doi:10.1080/14740338.2017.1323867

62. García-Abeijon P, Costa C, Taracido M, Herdeiro MT, Torre C, Figueiras A. Factors Associated with Underreporting of Adverse Drug Reactions by Health Care Professionals: A Systematic Review Update. Drug Saf. 2023;46(7):625–636. doi:10.1007/s40264-023-01302-7

63. Pariente A, Gregoire F, Fourrier-Reglat A, Haramburu F, Moore N. Impact of safety alerts on measures of disproportionality in spontaneous reporting databases: the notoriety bias. Drug Saf. 2007;30(10):891–898. doi:10.2165/00002018-200730100-00007

64. De Boissieu P, Kanagaratnam L, Abou Taam M, Roux M, Dramé M, Trenque T. Notoriety bias in a database of spontaneous reports: the example of osteonecrosis of the jaw under bisphosphonate therapy in the French national pharmacovigilance database. Pharmacoepidemiology and Drug. 2014;23(9):989–992. doi:10.1002/pds.3622

65. Hoffman KB, Demakas AR, Dimbil M, Tatonetti NP, Erdman CB. Stimulated reporting: the impact of US food and drug administration-issued alerts on the adverse event reporting system (FAERS). Drug Saf. 2014;37(11):971–980. doi:10.1007/s40264-014-0225-0

66. Fukazawa C, Hinomura Y, Kaneko M, Narukawa M. Significance of data mining in routine signal detection: Analysis based on the safety signals identified by the FDA. Pharmacoepidemiology and Drug. 2018;27(12):1402–1408. doi:10.1002/pds.4672

67. Dhodapkar MM, Shi X, Ramachandran R, Chen EM, Wallach JD, Ross JS. Characterization and corroboration of safety signals identified from the US Food and Drug Administration Adverse Event Reporting System, 2008-19: cross sectional study. BMJ. 2022;379:e071752. doi:10.1136/bmj-2022-071752

