## Supplemental Methods Text for "Patient-Level Risk Characterization of Drug-Associated Hidradenitis Suppurativa Using Machine Learning"

**eText. Methods Supplement**

**Disproportionality Analysis**

Analyses were restricted to primary suspect drugs with 3 or more HS reports. Drug-HS signals were quantified using reporting odds ratios (RORs) within strata defined by HS indication history, sex, and a combined total. For each drug-stratum combination, 2 × 2 tables compared HS vs non-HS outcomes across primary-suspect exposed vs unexposed reports.^20,27,28^ The ROR was calculated as (a × d)/(b × c), with Wald 95% CIs and 2-sided P values from z tests on log(ROR). Statistical significance was defined as P < .05, and reporting-odds-ratio P values were additionally adjusted for multiple comparisons using the Benjamini-Hochberg false discovery rate within each HS-indication-by-sex analysis stratum.

**Machine-Learning Model Development, Calibration, and Benchmarking**

Full computational specifications for the rare-event risk-modeling framework.

**1. Cohort, outcome, and candidate features**

Models were trained on the full tumor necrosis factor inhibitor (TNFi)-exposed FAERS population (N = 1 451 950 reports; HS defined by OMOP concept 37320281). Reports were split 70/30 with stratification on HS status into training (n = 1 016 365; 667 cases) and held-out test (n = 435 585; 286 cases) sets (seed = 123). The candidate feature set contained 62 routinely reported fields: TNFi agent identity, treatment indication, age, age-missingness, sex, and binary concomitant-medication and comorbidity indicators for disease context. All models used the identical set of selected features (below); no test-set information informed training, feature selection, or calibration.

**2. Class-imbalance weighting**

The HS-to-non-HS ratio was approximately 1523:1. Rather than resampling, each model received per-report weights, with minority (HS) reports up-weighted by min(N_majority/N_minority, 50). The weight cap of 50 prevented a small number of cases from dominating the loss and destabilizing probability estimates. The identical weighting function was applied within every cross-validation fold and to every algorithm that accepts sample weights.

**3. Feature selection**

Features were selected by 10-fold stratified cross-validation on the training set (seed = 456). Within each fold a random forest (500 trees, max_features = √p, min_samples_leaf = 5, fully grown trees, out-of-bag scoring, per-fold seed 1000 + fold) was fit and features ranked. Features selected in at least 5 of 10 folds were retained, yielding a parsimonious set of 29 candidate predictors used for all supervised models. Feature-selection stability (folds selected per feature) is reported in eFigure 10.

**4. Model development**

Five supervised algorithms representing complementary paradigms (penalized linear models, tree-based ensembles, and an interpretive-additive model) were trained on the 29 selected features with the weighting above. An unsupervised Isolation Forest served as a negative-control benchmark to establish whether HS reports behaved primarily as statistical outliers rather than a rather than a labeled reporting pattern recoverable by supervised learning. Hyperparameters are given in the Table below; all were prespecified (literature-standard values), not tuned on the test set. No supervised model was considered intrinsically preferred a priori. Because performance and identified predictors were highly concordant across approaches, the random forest was subsequently used as a representative model for visualization and interpretation. Penalized logistic regression served as the primary adjusted association model among the supervised approaches, while the separate multivariable logistic interaction model (see below) was used for formal indication-by-agent inference.

**eMethods Table. Prespecified hyperparameters for all models.**

| Model (library) | Primary Purpose | Key hyperparameters |
| --- | --- | --- |
| Random forest (scikit-learn) | Representative tree ensemble | 500 trees; max_features=√p; min_samples_leaf=5; max_depth=None (fully grown); OOB scoring; sample-weighted; seed 789 |
| Gradient-boosted trees / XGBoost | Gradient boosted ensemble | objective=binary:logistic; n_estimators=500; max_depth=6; learning_rate=0.05; subsample=0.8; colsample_bytree=0.8; min_child_weight=5; scale_pos_weight=min(ratio,50); seed 42 |
| Explainable boosting machine (interpret) | Interpretable additive model | Main effects only (interactions=0); default cyclic boosting; sample-weighted; seed 42 |
| Ridge (L2-penalized logistic) regression (scikit-learn) | Penalized linear benchmark | LogisticRegressionCV; penalty=L2; 10-fold CV; scoring=ROC-AUC; max_iter=1000; sample-weighted; seed 123 |
| Elastic-net logistic regression (scikit-learn) | Sparse linear benchmark | LogisticRegressionCV; solver=saga; l1_ratios=0.2/0.5/0.8; Cs=5; 5-fold CV; scoring=average precision; max_iter=3000; sample-weighted; seed 123 |
| Isolation forest (scikit-learn) | Anomaly-detection benchmark | n_estimators=300; max_samples=auto; contamination=auto; anomaly score = −score_samples; seed 42 |

*√p, square root of the feature count; ratio, majority-to-minority class ratio; OOB, out-of-bag.*

**5. Probability calibration**

Because the event rate was 0.07%, probabilities were calibrated by out-of-fold (OOF) isotonic regression to prevent leakage: OOF training-set predictions were generated by stratified cross-validation, an isotonic map was fit on those OOF predictions, and the map was applied to the test predictions. The reference random forest used 5-fold OOF isotonic calibration (seed = 800); XGBoost used 5-fold OOF isotonic (seed = 900); the remaining supervised models used a shared 3-fold OOF isotonic helper (seeds 202–203).

Predicted probabilities were calibrated by out-of-fold isotonic regression.^31–35^ For the reference model, four calibration methods were compared: raw, isotonic, Platt scaling (on a class-balanced subsample of ≤50 000 controls; unpenalized logistic link), and three-parameter beta calibration (betacal, parameters=“abm”). Isotonic was selected to serve as primary calibration method based on Brier score and predicted-to-observed ratio (eTable 14; eFigure 9). Because isotonic calibration is monotonic, it leaves rank-based metrics (AUPRC, top-percentile enrichment) unchanged; calibrated probabilities were therefore used only for calibration metrics and not for comparative evaluation of enrichment or discrimination.

**6. Performance evaluation and benchmarking**

All models were evaluated on the single held-out test set. Because discrimination thresholds are uninformative at this event rate, performance was led by (a) the area under the precision-recall curve (AUPRC), with 95% CIs from 1000 bootstrap resamples, and (b) enrichment across predicted risk strata—the fold-increase in HS rate and the percentage of all HS cases captured within the highest-risk 1%, 5%, and 10% of ranked reports, with bootstrap CIs. Secondary metrics were the area under the receiver operating characteristic curve (AUROC) and calibration (Brier score, predicted-to-observed ratio, expected and maximum calibration error using both uniform and quantile bins). Models performance was compared on raw scores and in a single unified table (eTable 20); per-model enrichment detail appears in Figure 3C-D. The random forest was compared against the logistic-regression benchmark by a bootstrap AUROC difference test (2000 resamples).

**7. Model interpretation**

The reference random forest was explained with TreeSHAP computed on 2000 randomly sampled test reports (seed = 101), summarized by mean absolute SHAP value. Permutation importance was computed on the test set (10 repeats; seed = 792; scored by AUROC decrease) as an independent check (eTable 19). The explainable boosting machine provided per-feature shape functions and global term importances, plus local additive explanations for a representative correctly ranked (true-positive) versus missed (false-negative) report (Figure 4). Candidate pairwise feature interactions were screened by the correlation of per-report SHAP values (eTable 21); this proxy flags co-directional effects and does not by itself isolate synergistic interaction, motivating the formal analysis below.

To compare feature prioritization across supervised models, permutation importance was calculated on the held-out test set (10 repeats; seed = 42; an identical test subsample comprising all HS cases plus up to 80 000 randomly sampled controls was used for every model) as the decrease in AUROC after feature shuffling, and normalized within each model. Concordance was quantified using pairwise Spearman correlations (ties broken by average ranks) across all retained features and within the union of each model pair’s top 10 features; overlap among the top 5 and top 10 features was also calculated.

**8. Formal interaction analysis (inferential)**

To assess effect modification independent of the predictive models, we fit a logistic regression for the indication × TNFi-agent interaction on the full cohort (statsmodels), adjusting for age group, sex, and the 15 concomitant medications with the highest mean absolute SHAP values (missingness modeled as its own level, avoiding imputation) to limit dimensionality.

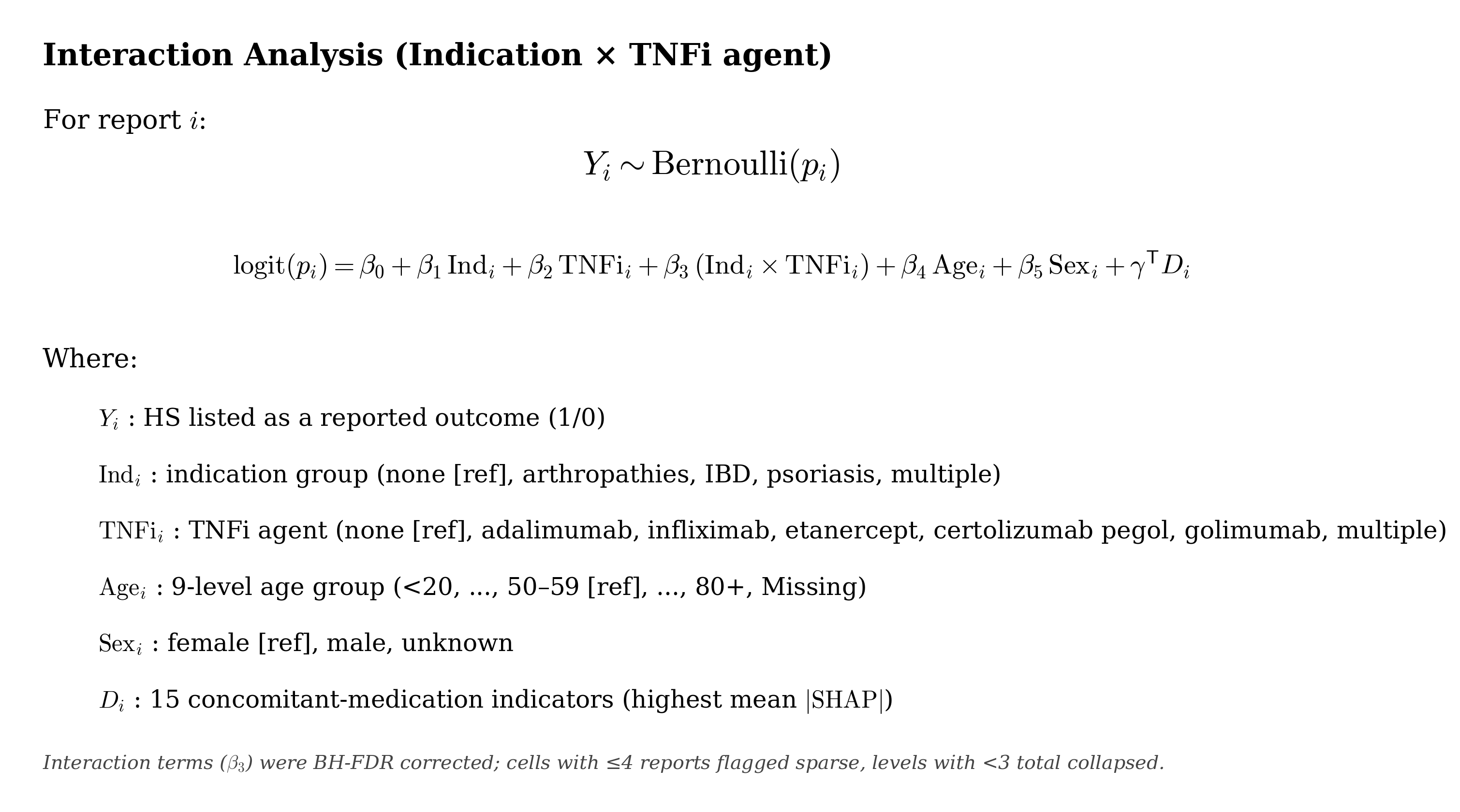

Interaction-term P values were Benjamini-Hochberg FDR-corrected (full output, eTable 8; interaction odds ratios, eTable 11); sparse cells (≤4 reports) were flagged and excluded from primary inference, and category levels with fewer than 3 total reports were collapsed (eTable 10). Adjusted HS probabilities by indication and agent were obtained by marginal standardization (g-computation; eTable 9).

**9. Software and reproducibility**

Analyses used Python 3.12.13 with scikit-learn v1.6.1 (random forest, penalized and elastic-net logistic regression, isotonic regression, permutation importance, isolation forest), XGBoost v3.3.0 (gradient-boosted trees), interpret v0.7.8 (explainable boosting machine), shap v0.52.0 (TreeSHAP), betacal (beta calibration), statsmodels v0.14.6 (interaction regression, Benjamini-Hochberg FDR), and SciPy/NumPy/pandas. Gemini 3.1 Pro was used for code generation; all code and outputs were reviewed and validated by the authors. All random seeds were fixed as specified above and in the table to permit exact reproduction; the full analysis notebook is available with the data-availability materials.
