## Supplemental Figures for "Patient-Level Risk Characterization of Drug-Associated Hidradenitis Suppurativa Using Machine Learning"

**eFigure 1.** Study design and analytic cohorts

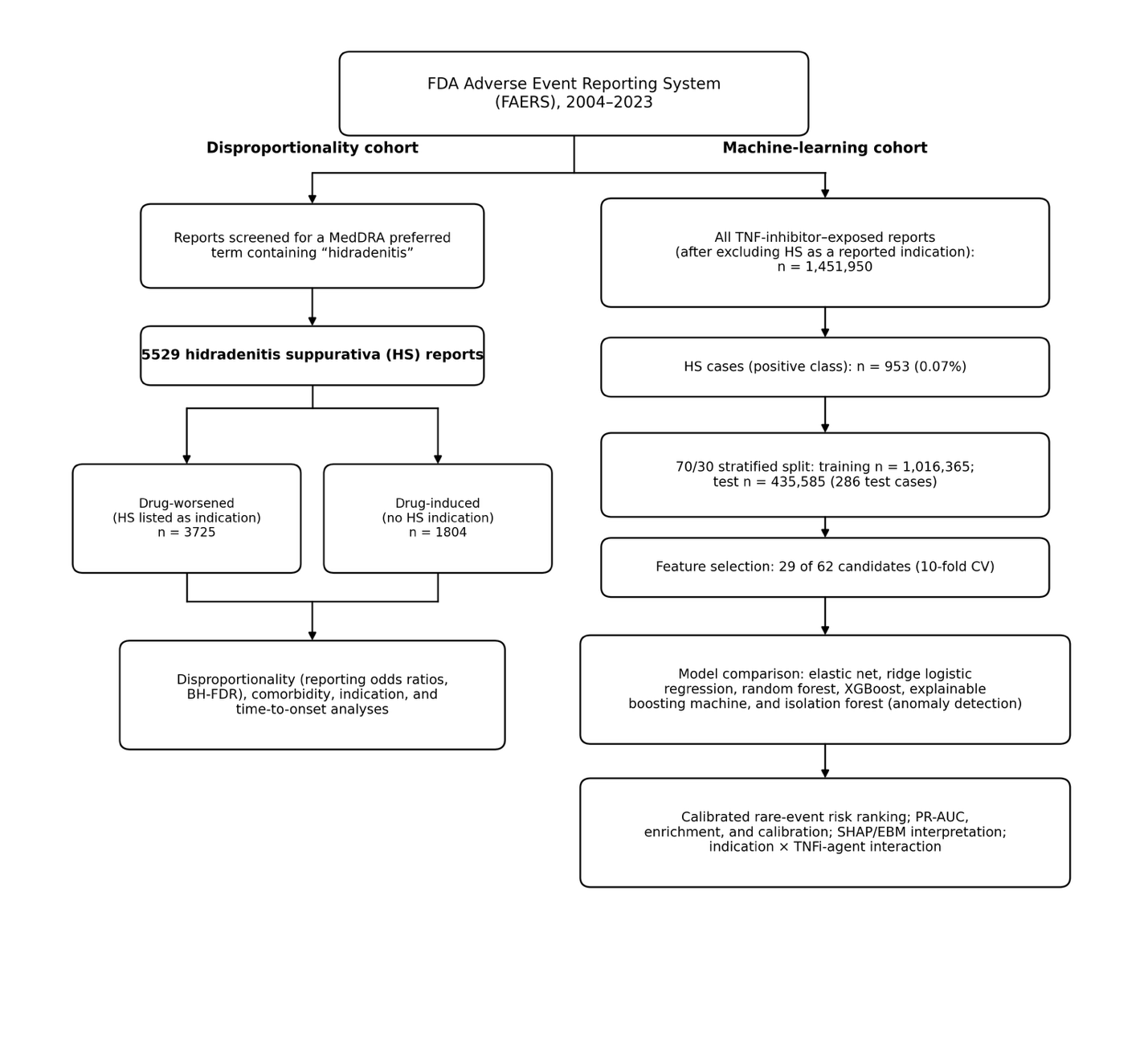

FAERS reports (2004–2023) were screened for a MedDRA preferred term containing "hidradenitis," yielding 5,529 hidradenitis suppurativa (HS) reports stratified into drug-worsened (DW, n=3,725) and drug-induced (DI, n=1,804) groups for disproportionality, comorbidity, indication, and time-to-onset analyses. In parallel, the full TNF-inhibitor (TNFi)–exposed population (n=1,451,950, after excluding HS as a reported indication) was used to train and compare machine-learning risk models, with TNFi-exposed HS cases (n=953) as the positive class and remaining TNFi-exposed reports as controls, followed by an indication × TNFi-agent interaction analysis.

Abbreviations: FAERS, FDA Adverse Event Reporting System; HS, hidradenitis suppurativa; MedDRA, Medical Dictionary for Regulatory Activities; TNFi, tumor necrosis factor inhibitor; DW, drug-worsened; DI, drug-induced.

**eFigure 2.** Hidradenitis Suppurativa (HS) reports by year and age, stratified by prior history of HS

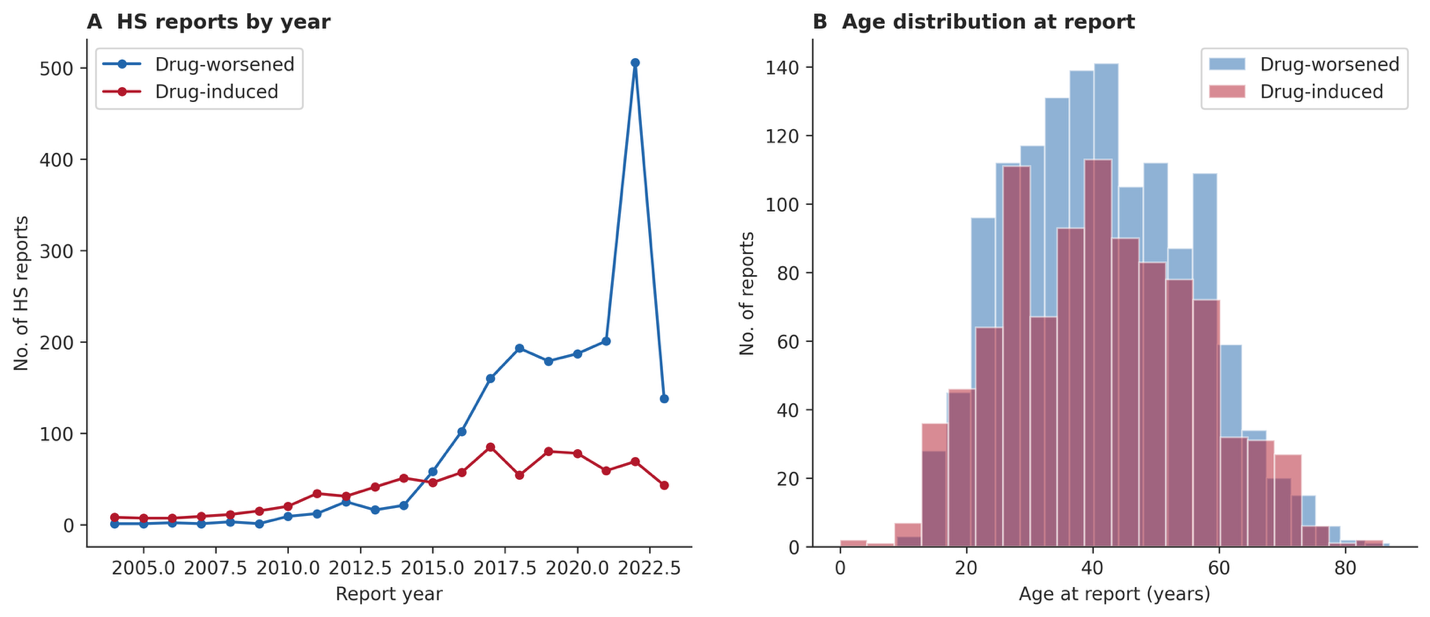

Annual HS outcome report counts (A) and age distribution at report (B), shown separately for reports with HS listed as an indication (drug-worsened, DW) and reports without HS listed as an indication (drug-induced, DI).

Abbreviations: HS, hidradenitis suppurativa; DW, drug-worsened; DI, drug-induced.

**eFigure 3.** Top indications for receiving primary suspect drug associated with HS outcome
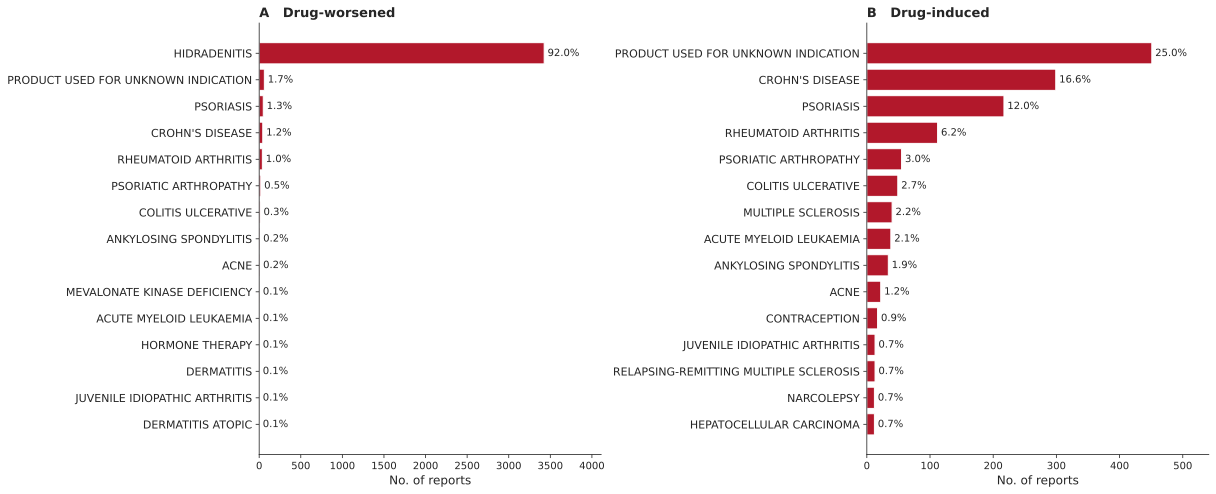

Most frequently reported indications for the primary suspect drug among reports with HS listed as an indication (drug-worsened, DW, panel A) and among reports without HS listed as an indication (drug-induced, DI, panel B). Reports with missing or “unknown” indication were excluded.

Abbreviations: HS, hidradenitis suppurativa; DW, drug-worsened; DI, drug-induced.

**eFigure 4.** Indication-derived disease-context profiles among HS outcome reports.

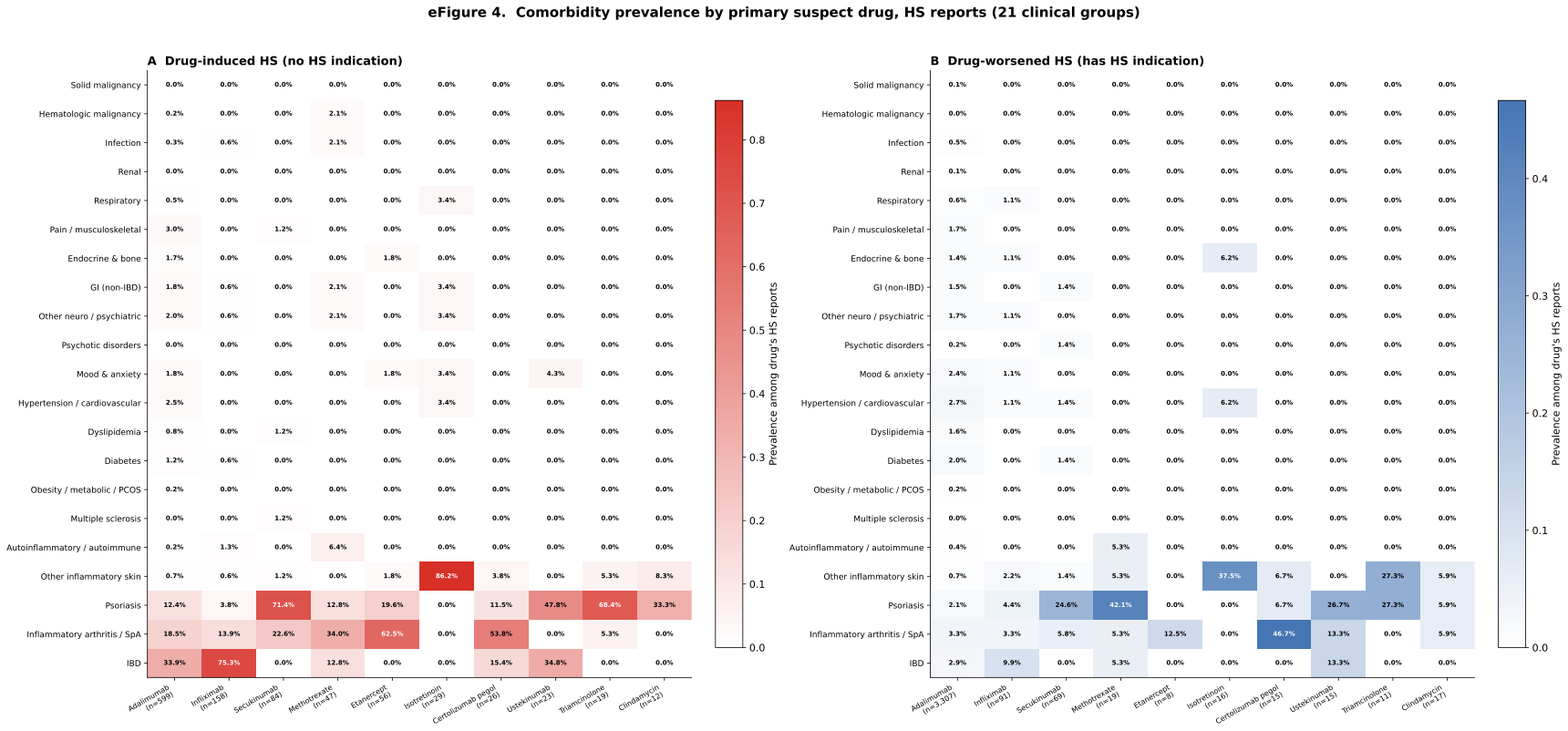

Indication-derived disease context among reports without HS listed as an indication (drug-induced, DI; A) and reports with HS listed as an indication (drug-worsened, DW; B), restricted to selected primary suspect drugs. Tiles show the proportion of HS outcome reports carrying each comorbidity flag, annotated as percentages (scale fixed 0–100%).

Abbreviations: HS, hidradenitis suppurativa; DI, drug-induced; DW, drug-worsened.

**eFigure 5.** Mirrored HS Case-Count Signal Plot for Top Drugs by Indication History

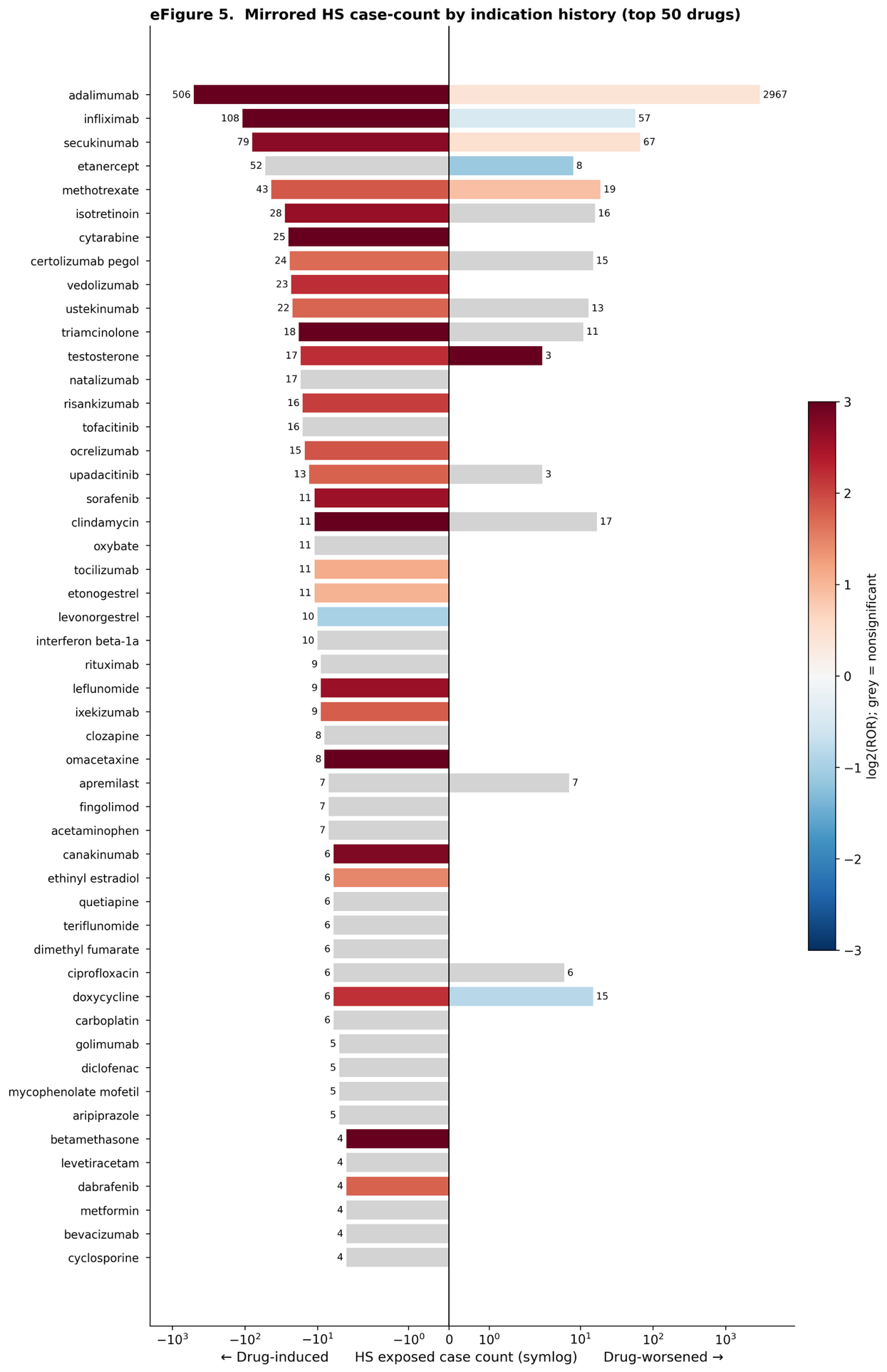

Mirrored bar plot showing HS exposed case counts for the top 50 drugs ranked by HS case count in the drug-induced (no HS indication) stratum. Left bars represent counts among reports without HS listed as an indication (drug-induced); right bars represent counts with HS listed as an indication (drug-worsened). Bar color encodes disproportionality strength (reporting odds ratio [ROR]) for statistically significant signals (P < .05), centered at ROR = 1 (white) using a log-transformed scale (blue, ROR < 1; red, ROR > 1). Gray bars indicate nonsignificant signals. Counts are displayed on a pseudo-log scale.

Abbreviations: HS, hidradenitis suppurativa; ROR, reporting odds ratio; DI, drug-induced; DW, drug-worsened.

**eFigure 6.** Time-to-Onset Profiles by Drug Class for Paradoxical HS

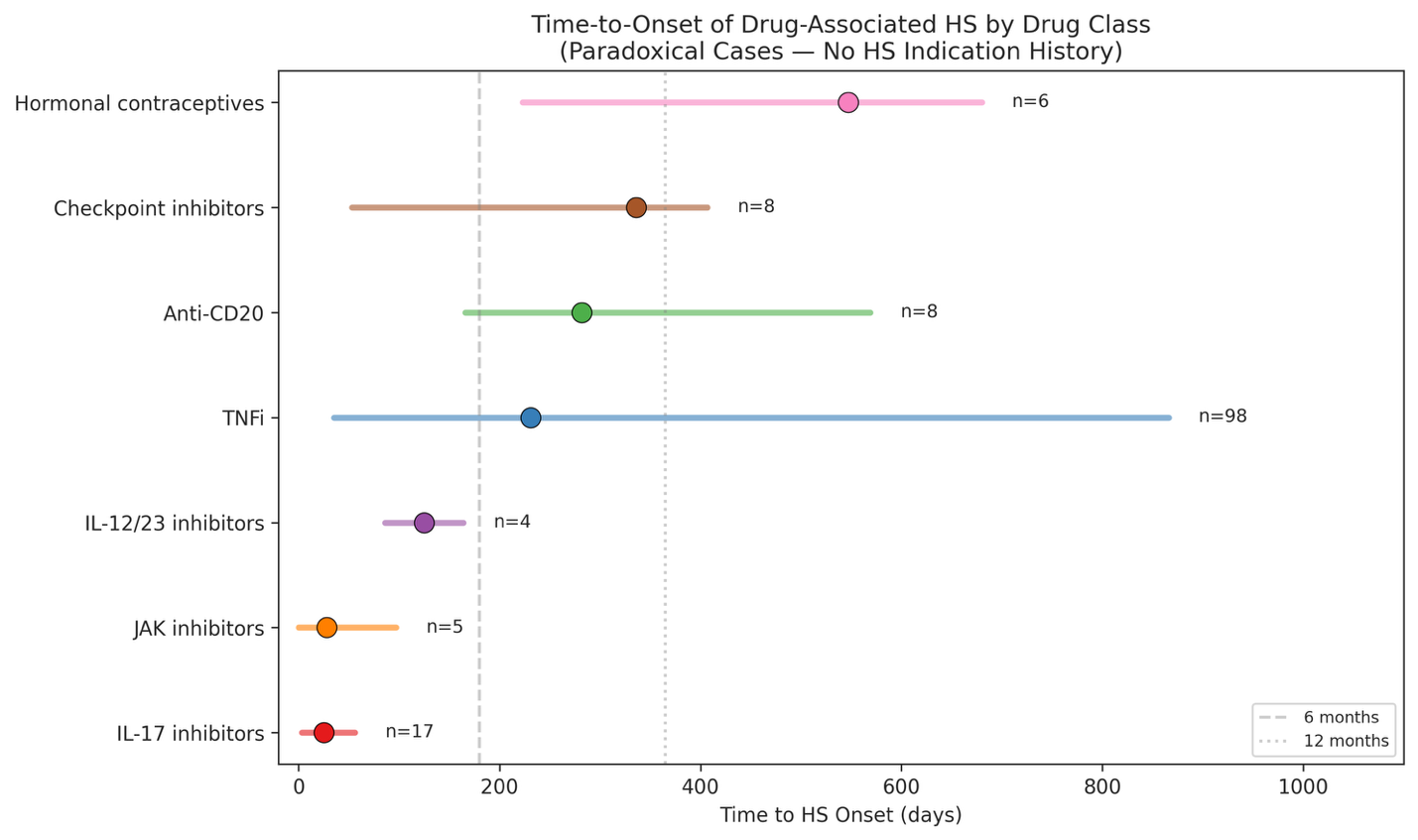

Summary of time-to-onset among paradoxical (no HS history) reports by drug class. Data presented as median days with interquartile range. TNFi (n = 98) show gradual onset (median, 231 days), while IL-17i (n = 17) demonstrate rapid onset (median, 25 days). JAKi (n = 5; median, 28 days), anti-CD20 agents (n = 8; median, 282 days), and checkpoint inhibitors (n = 8; median, 336 days) are also shown. Top contributing agent listed for each class.

Abbreviations: HS, hidradenitis suppurativa; TNFi, tumor necrosis factor inhibitor; IL-17i, interleukin-17 inhibitor.

**eFigure 7.** Sex-Stratified Volcano Plot of Risk Signals (Drug-Induced HS)

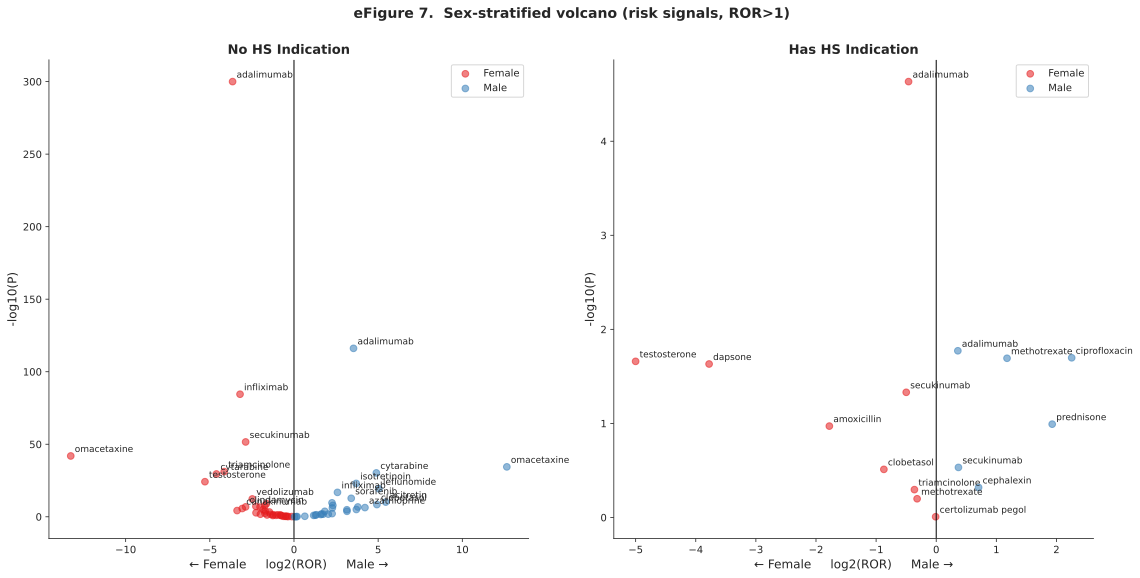

Volcano plots of sex-stratified disproportionality signals restricted to risk signals (reporting odds ratio [ROR] > 1), faceted by HS indication history. The x-axis represents log2(ROR) and the y-axis –log10(P). Female signals are mirrored to the left and male signals to the right. The 10 most significant drugs are labeled within each sex-by-history stratum.

Abbreviations: HS, hidradenitis suppurativa; ROR, reporting odds ratio.

**eFigure 8. Explainable boosting machine importances and isolation-forest anomaly scores**

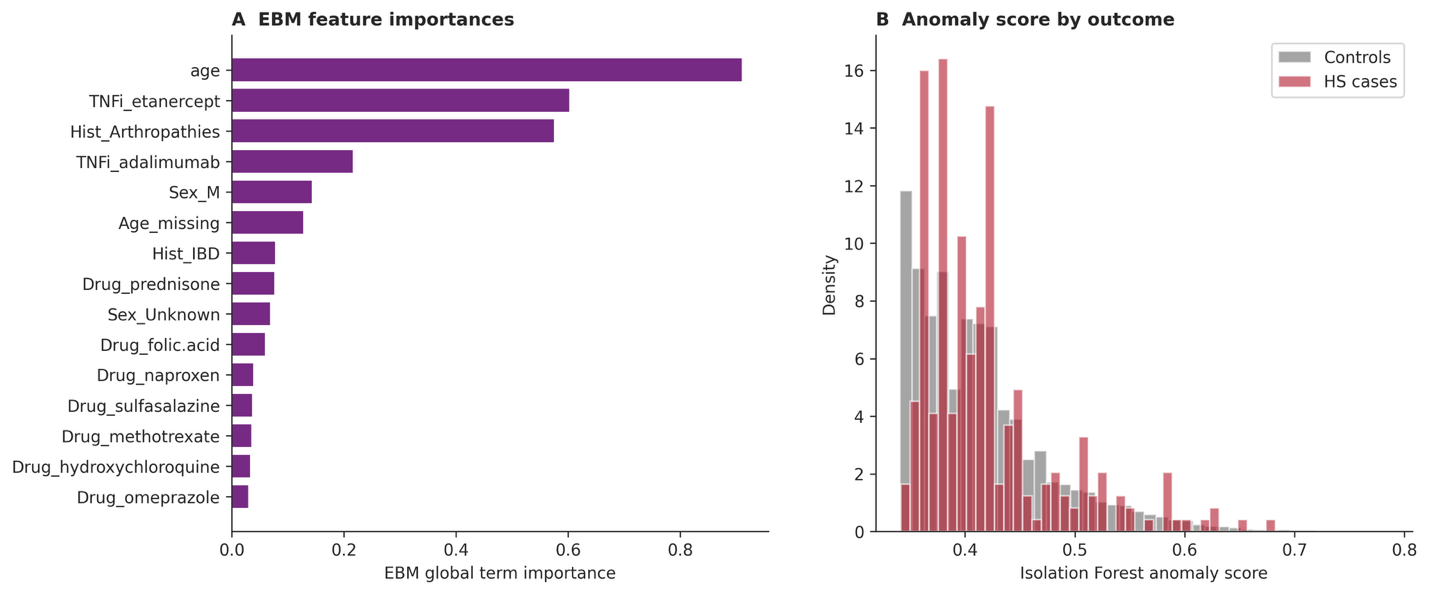

(A) Global term importances from the explainable boosting machine (EBM). (B) Distribution of unsupervised isolation-forest anomaly scores among HS cases versus controls, showing substantial overlap consistent with the near-chance discrimination of anomaly detection for this outcome.

Abbreviations: HS, hidradenitis suppurativa; EBM, explainable boosting machine.

**eFigure 9. Calibration of isotonic-scaled probabilities across supervised models**

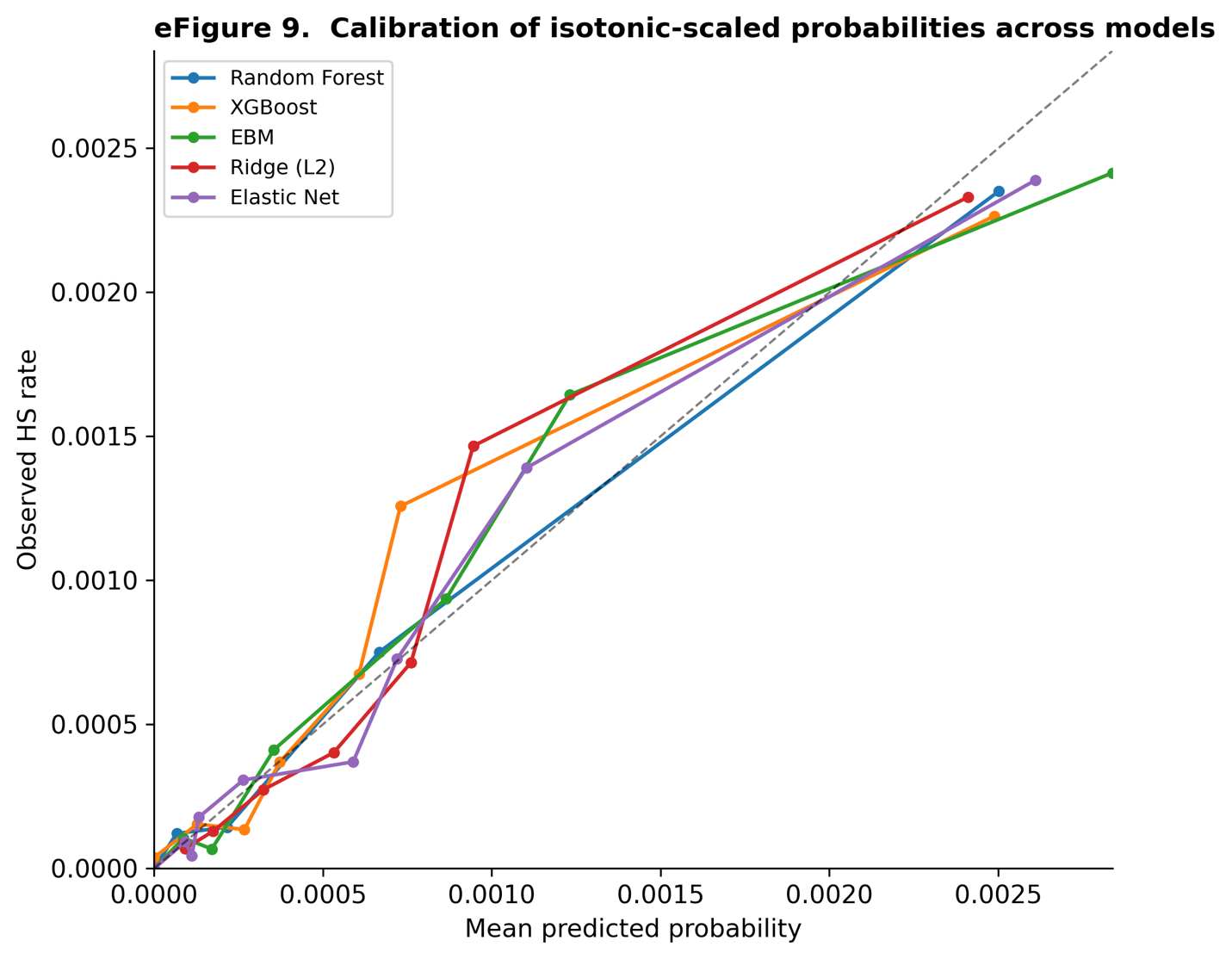

Reliability curves (observed vs mean predicted probability) for the supervised models after out-of-fold isotonic calibration, zoomed to the event-rate region. Points on the diagonal indicate accurate calibration.

Abbreviations: HS, hidradenitis suppurativa.

**eFigure 10. Cross-model feature-importance concordance.**
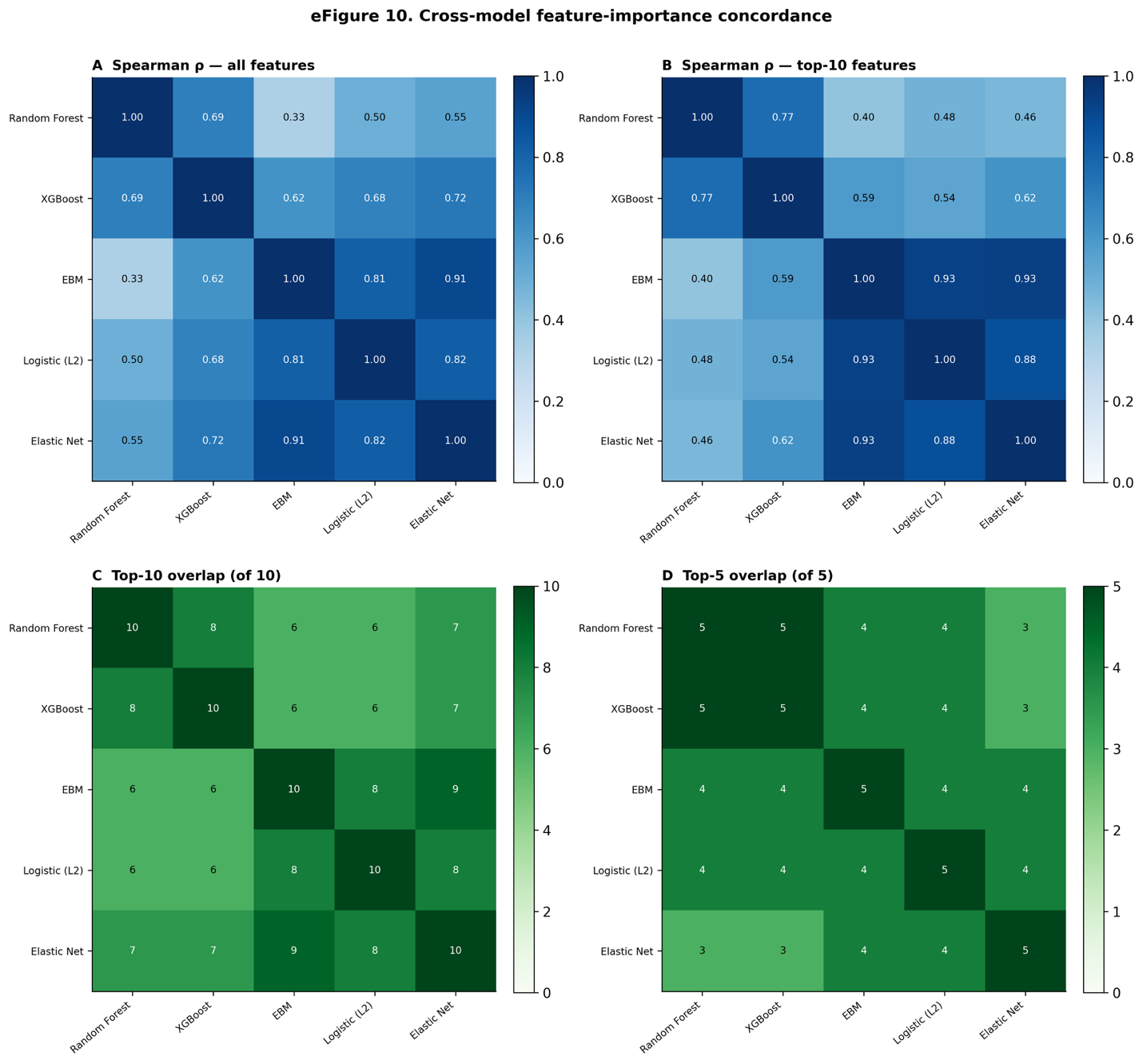

For each supervised model, permutation importance was computed for the 29 retained features on the held-out test set and features were ranked. **A**, Pairwise Spearman correlation of rankings across all features. **B**, Spearman correlation restricted to the union of each model's top-10 features. **C**, Number of shared features among each model pair's top 10. **D**, Shared features among each pair's top 5.
